# Facilitators and barriers to SGLT2i and GLP1a prescribing in Northern Ontario: a qualitative interview study

**DOI:** 10.1101/2025.10.27.25338651

**Authors:** Patricia Olar, Carolyn Steele Gray, Tamara Van Bakel, Joseph Benjamin, Michael Fralick

## Abstract

**Background:** One in three adults in Ontario, Canada has type 2 diabetes, obesity, heart failure, or chronic kidney disease, and the prevalence is even higher in Northern Ontario. Sodium glucose co-transporter 2 inhibitors (SGLT2i) and glucagon-like peptide-1 analogues (GLP1a) are highly effective medications to treat these conditions, but prescribing rates in Northern Ontario are low. This study aimed to explore the facilitators and barriers to SGLT2i and GLP1a prescribing for adults living with and without diabetes in Northern Ontario.

**Methods:** We conducted virtual, semistructured interviews of clinicians (i.e., physicians, nurse practitioners, resident physicians) working in Northern Ontario, Canada between July 2024 and November 2024. Interview transcripts were thematically coded into categories based on the Theoretical Domains Framework (TDF). Findings were classified as either barriers or facilitators, and then grouped to identify major subthemes within the data. Subthemes were then further aggregated into themes and mapped onto the Capability, Opportunity, Motivation-Behaviour (COM-B) model for behaviour change.

**Results:** We interviewed 25 clinicians, including eight physicians, eight resident physicians, and nine nurse practitioners caring for adults in Northern Ontario. Twenty-two of the interviews were held one-on-one and one was held as a co-interview with three participants. We identified five main barriers and five main facilitators to SGLT2i and GLP1a prescribing. The major barriers included: limited access to medications, patient challenges and competing demands, lack of familiarity, clinical identity, and prescribing inertia. Limited access to medications was a prominent theme with nested subthemes of high cost of medications for patients and insufficient compassionate drug programs to cover these costs. The major facilitators included: role as a clinician that follows the data, belief that SGLT2i/GLP1a use will improve patient outcomes, clinicians’ perceptions of patient openness to these drugs, comfort prescribing, and system and colleague supports.

**Conclusion:** Our findings provide useful insights to inform knowledge translation initiatives aimed at increasing the uptake of SGLT2i and GLP1a in Northern Ontario.

## INTRODUCTION

Over the past decade, research on sodium glucose co-transporter 2 inhibitors (SGLT2i) and glucagon-like peptide 1 analogues (GLP1a) has rapidly established a strong evidence base for the broad benefit profiles of these medications in adults with chronic conditions, including type 2 diabetes, obesity, heart failure, and chronic kidney disease (CKD). All available SGLT2i are oral medications. GLP1a are available as both oral medications and subcutaneous injections; however, at the time of our study, the most commonly prescribed GLP1a (semaglutide) was only available in Canada as an injection.

Multiple randomized controlled trials have demonstrated that SGLT2i and GLP1a not only improve glycemic control but also improve cardiovascular and renal outcomes.^1,2^ Specifically, SGLT2i and GLP1a reduce the relative risk of adverse cardiovascular events (i.e., myocardial infarction, stroke, and cardiovascular death) by 10% and 14%, respectively; heart failure by 23% and 11%; and progression to end-stage renal failure by 38% and 21%.^1–5^ As a result, both drug classes are now recommended as second-line therapies for adults with type 2 diabetes. Notably, the evidence also indicates that the cardiorenal and weight loss benefits of these agents applies regardless of whether a person has diabetes, which has led to the inclusion of SGLT2i in Canadian clinical guidelines for heart failure and chronic kidney disease.^6–11^

Based on the prevalence of type 2 diabetes, obesity, heart failure, and chronic kidney disease in Ontario, approximately 35% of adults stand to benefit from SGLT2i and GLP1a.^12–19^ A recent study of over 1.2 million hospitalizations across Ontario identified that approximately 31% of patients had type 2 diabetes and 26% had obesity, but use of GLP1a was uncommon (<1%).^20^ SGLT2i use in this population was also low (5%), which aligns with previous studies.^20, 21^ Racial and ethnic minority groups and patients living in underserviced areas face a disproportionately high burden of type 2 diabetes and its related complications, yet are less likely to be treated with an SGLT2i or a GLP1a.^22–29^ This treatment gap is especially pronounced among adults living in Northern Ontario, where rates of diabetes can be as high as 32% in northern and rural First Nations communities, compared to an average prevalence of less than 10% province-wide.^30^

To gain insight on why prescribing rates of SGLT2i and GLP1a are low, we conducted a qualitative study interviewing clinicians who care for adults in Northern Ontario. Our objective was to identify facilitators and barriers to SGLT2i and GLP1a prescribing for adults living with and without diabetes.

## METHODS

### Study Design and Setting

Our qualitative work followed a pragmatic descriptive approach to data collection and analysis. This approach is considered suitable for pragmatic qualitative studies seeking to have real-world and clinical application.^31^ The study was conducted with clinicians working in Northern Ontario.

### Theoretical Approach

Our work was fundamentally grounded in the Theoretical Domains Framework (TDF) and the Capability, Opportunity, Motivation-Behaviour (COM-B) model for behaviour change.^32, 33^ Both frameworks are tools used in implementation science to understand the factors influencing behaviour change, particularly in healthcare settings. The TDF outlines 14 domains which may underlie behaviour change, while the COM-B model links behaviour change to either capability (one’s physical or psychological abilities), opportunity (social or environmental factors), and motivation (both automatic and reflective processes). As the COM-B model provides an overarching structure for understanding behaviour and the TDF offers a more granular view of underlying determinants, using both frameworks together allowed for a layered exploration of clinicians’ prescribing practices. The manner in which we used these frameworks in our data analysis are described below.

### Researcher Reflexivity

The research team consists of health services researchers, clinician scientists, and trainees with diverse training across multiple disciplines, including implementation and behaviour science and qualitative methods. Notably, program lead MF is a clinician scientist who is actively engaged in this clinical work. He has expertise in SGLT2i and GLP1a and has been a locum physician in Northern Ontario for the past nine years. The study was informed by his experiences working in Northern Ontario, where he observed that both classes of medications were prescribed relatively rarely. The research team met regularly to engage in reflexive discussion around data and analysis, reflecting on where previous experience and training may influence analysis and interpretation.

### Ethics

The Mount Sinai Hospital Research Ethics Board of Sinai Health System gave ethical approval for this work.

### Participants

Our intended sample size was 30-45 clinicians, including approximately 10-15 staff physicians, 10-15 resident physicians, and 10-15 nurse practitioners working in both inpatient and outpatient settings. This sample size is consistent with qualitative studies of this type to achieve sufficient thematic saturation and conceptual depth.^34,35^ We used broad inclusion criteria to capture insights from clinicians with experiences in diverse clinical contexts who collectively share the responsibility of caring for adults with chronic diseases in Northern Ontario. We recruited interview participants through our study team’s networks, cold emailing, posts on X and LinkedIn, and snowball sampling.

### Procedure

We obtained and documented verbal consent from all participants immediately prior to beginning each interview. All interviews were hosted on Zoom and led by PO, excluding one which was led by JB. Our interview guide was developed by MF and PO, with guidance from CSG, who has expertise in qualitative research (see our full interview guide in **Appendix A**). Interviews were audio recorded (video was turned off), and the audio recordings were then manually transcribed verbatim by PO. Interview transcripts maintained participant anonymity. Following transcription, all recorded files were deleted. All participants were offered a gift card honorarium for their participation in the study.

### Data Analysis

To analyze the data, we followed an abductive analytic approach. First, we conducted a thematic analysis of each transcript.^36^ PO reviewed each transcript and deductively assigned codes to salient statements from each interview based on our codebook. Early in the coding process, PO reviewed two coded transcripts in a meeting with CSG and MF to discuss whether code assignment adequately matched the sentiment expressed by the interviewee and to establish the first iteration of the codebook. Our codebook was fundamentally grounded in the TDF. We expanded upon the 14 domains outlined in the TDF in an iterative process to include sub-groups specific to our data as new ideas were articulated in the interviews. For example, under the domain Social Influences, we differentiated influences from preceptors, colleagues, patients, and the media as different sub-groups (see our full codebook in **Appendix B**). We coded statements to multiple TDF domains if more than one domain was deemed to be applicable.

Next, we conducted inductive coding in two stages: first, to interpret and classify statements as either barriers or facilitators, and second, to group similar statements together and form subthemes. Subthemes were kept linked to their relevant TDF domains. We then clustered subthemes into themes and mapped the themes onto the COM-B model. Our process of identifying and defining themes and subthemes occurred both asynchronously by PO and synchronously in team meetings held between PO, MF, CSG and TVB. Our themes, subthemes, definitions, and some exemplary quotes are outlined comprehensively in **Appendix C**. Analytic rigour was enhanced in our study through the following measures: (1) we had multiple researchers assist in collating subthemes into themes, (2) we maintained an audit trail of our analysis at every step, and (3) we followed the Standards for Reporting Qualitative Research guidelines.^37^

## RESULTS

We interviewed 25 clinicians, including eight physicians, eight resident physicians, and nine nurse practitioners, caring for adults in Northern Ontario. Twenty-two interviews were conducted one-on-one and one interview was held as a co-interview with three nurse practitioners. Seventeen clinicians worked in outpatient settings (including family medicine clinics, nurse practitioner-led clinics, walk-in clinics, diabetes education centres, and emergency departments), four worked in inpatient settings, and four worked in both inpatient and outpatient settings. The average audio recording length was 21:00 minutes (range 8:48-29:01 minutes). Interviews were conducted between July 2024 and November 2024. Data crystallization was achieved in the last five interviews conducted. We defined data crystallization as the point at which our iterative analysis across interviews revealed consistent and well-integrated themes, indicating that our dataset had achieved conceptual depth and that further data collection was unlikely to provide novel insights.

We identified 28 subthemes that mapped to 12 TDF domains and encompassed all three components of the COM-B model. We assembled these subthemes into five main barriers and five main facilitators to SGLT2i and GLP1a prescribing. Our pool of interviewed clinicians generally described familiarity with and common use of SGLT2i and GLP1a; however, our interviews still emphasized several key barriers to their uptake. Important facilitators of SGLT2i and GLP1a prescribing also emerged through the interviews. See **Table 1** for the main barriers and facilitators and **Appendix C** for a comprehensive table including our themes, subthemes, definitions, and some quotes.

**Table 1.** Barriers and facilitators to SGLT2i and GLP1a prescribing mapped to the TDF domains.

| Themes and Subthemes | TDF Domains |
| --- | --- |
| **Barriers** |  |
| **Limited access to medications** |  |
| Cost of the medications to patients | Environmental context & resources |
| Insufficiency of Compassionate Drug Programs | Environmental context & resources |
| Periodic drug shortages | Environmental context & resources |
| Clinician shortage | Environmental context & resources |
| **Patient challenges and competing demands** |  |
| Medication averse | Social influences |
| Fear of side effects | Social influences |
| Medication non-adherence and appointment no-show | Social influences |
| **Lack of familiarity** |  |
| Not enough experience with patients using the medications | Skill; Beliefs about consequences; Emotions |
| Knowledge gaps | Knowledge; Skill; Memory, attention, and decision processes |
| Lack of confidence | Skill; Belief about capabilities |
| **Clinical identity** |  |
| Risk tolerance | Social/professional role and identity; Beliefs about consequences; Intention; Memory, attention, and decision processes; Emotion |
| Perceived scope of practice | Social/professional role and identity |
| Hesitancy accepting GLP1a for weight loss | Social/professional role and identity; Social influences |
| Clinical practice guideline-directed | Social/professional role and identity |
| **Prescribing inertia** |  |
| Routine patterns of prescribing | Behavioural regulation; Memory, attention, and decision processes |
| **Facilitators** |  |
| **Role as a clinician that follows the data** |  |
| Respecting clinical guidelines | Social/professional role and identity |
| Guided by evidence | Social/professional role and identity; Intention |
| **Belief that SGLT2i/GLP1a will improve patient outcomes** |  |
| Desire to offer their patients medications with strong benefit profiles | Intention |
| Optimism that medication use will improve health outcomes | Optimism |
| **Clinicians' perceptions of patient openness** |  |
| Patients express desire for weight loss | Social influences |
| Patients understand second-line treatment may be needed | Social influences |
| **Comfort Prescribing** |  |
| Experience builds skill and confidence | Skill; Beliefs about capabilities |
| Prevalence of SGLT2i/GLP1a builds comfort prescribing | Knowledge; Skill; Social influences; Emotion |
| SGLT2i are easy to prescribe | Skill; Memory, attention, and decision processes |
| SGLT2i/GLP1a are backed by good data | Knowledge; Beliefs about capabilities |
| Prescribing is important to scope of practice | Social/professional role and identity |
| **System and Colleague Supports** |  |
| Expanding eligibility criteria for drug coverage | Environmental context & resources |
| Colleagues motivate ongoing learning | Social influences |

Abbreviations: GLP1a, glucagon-like peptide 1 analogues; SGLT2i, sodium glucose co-transporter 2 inhibitors.

### Barriers

#### Theme 1: Limited access to medications (Opportunity)

Clinicians described being limited in their ability to prescribe SGLT2i and GLP1a by several system-level barriers. Clinicians framed these barriers as interfering with patient care by reducing patient access to the medications even when prescribers and patients alike see the benefit of starting on these agents. From clinicians’ perspectives, the four major obstacles encountered are as follows. First, SGLT2i and GLP1a are both expensive drug classes for patients who lack adequate drug insurance and for those who do not meet strict Limited Use code criteria. Some clinicians reported that up to 40-50% of their patient population includes adults who are unemployed or otherwise lack sufficient private drug coverage to pay for a GLP1 medication––particularly semaglutide––out of pocket. In these scenarios, clinicians are forced to consider more affordable alternatives, including sulfonylureas.

> *“…definitely in Northern Ontario, there’s a lot of cost concerns. A lot of patients are…they’re sort of this in-between, where they don’t have [Ontario Disability Support Program funding] or they’re on Ontario Works and they don’t have medical coverage. But they’re not at that point where they’re over 65 and get [Ontario Drug Benefit] coverage, and they can’t afford their meds. So, they’ll take them or they won’t take them, or they’ll take them for a month and then they don’t take them..The other [option] is going for cheaper versions, like going for medications we wouldn’t normally prescribe but doing that because they’re cheaper.”*

While support programs to improve medication access exist (such as the Ontario Drug Benefit Plan and Compassionate Drug Programs), participants noted limitations. Notably, among patients without diabetes, even those over age 65 who qualify for the Ontario Drug Benefit plan face a roadblock: semaglutide is listed under a Limited Use code that restricts clinicians from prescribing it for weight loss unless the patient has a type 2 diabetes diagnosis and has failed to tolerate metformin.

Second, two clinicians who have turned to Compassionate Drug Programs to provide SGLT2i or GLP1a for their uninsured patients described these programs as inadequate for patient needs––mainly because medication supply is delayed month-to-month and terminates after only one year. These shortcomings limit the utility of such programs for long-term patient therapy.

A third obstacle clinicians described were drug shortages in Northern Ontario, another key example of missed opportunity to prescribe SGLT2i or GLP1a, even when clinicians are motivated to do so. This shortage was also apparent globally, related to a demand for semaglutide that outpaced supply.

Finally, given the high prevalence of chronic illness in Northern Ontario and the region’s relative shortage of clinicians, some clinicians described an insufficiency of the human capital needed to provide the community resources, patient education, and health care required to manage life-long conditions like type 2 diabetes. Many patients are thus not optimized on newer therapies like SGLT2i and GLP1a simply because they lack consistent access to primary care providers, nurse practitioners, or specialists who can initiate and monitor these treatments. As a result, the main opportunity to prescribe these medications often arises in acute care encounters such as emergency department visits or hospitalizations when patients present with complications or exacerbations.

> *“…because there are so few family doctors in Northern Ontario or even endocrinologists or anyone who can help manage those things or monitor the diabetes…that some people aren’t optimized because they don’t have primary care to monitor or even suggest, like some people could have been like, I don’t even know…‘I’m on metformin and I have been but I lost my family doctor 10 years ago’ and so they may not be optimized. People come into emerg all the time without a family doctor and they’re admitted to hospital, don’t have a family doctor, and while they’re in hospital, that’s when we try our best as an inpatient service to optimize their diabetes medications or get them the proper referrals to an internal medicine clinic or an endocrinologist so that they can be optimized.”*

#### Theme 2: Patient challenges and competing demands (Opportunity)

In several cases, clinicians reported being restricted from prescribing an SGLT2i or a GLP1a by patients themselves. Many clinicians cited patient refusal for reasons including resistance to increasing one’s pill burden, reluctance to break status quo once stable glycemic control had been achieved on other antihyperglycemic medications, fear of GLP1a injections, fear of the medications’ side effect profiles, and discomfort with GLP1a being so highly stigmatized in the media. Among those who rejected SGLT2i were women who did not want to risk developing urinary frequency or recurrent urogenital infections, especially if they had previously had a urinary tract or yeast infection.

> *“I have a number of women who when you tell them it might increase the risk of yeast infection, they want to run away from it….I’ve had patients tell me: ‘I’ve had yeast infections. There’s no way I ever want a yeast infection and I don’t want empagliflozin [SGLT2i].’”*

For some patients who were initially prescribed an SGLT2i or GLP1a, the occurrence of side effects led many to prefer terminating the prescription shortly thereafter.

Some clinicians also reported major problems with medication non-adherence and appointment no-shows in their patient populations. The reasons for both are multifactorial and highlight challenges and competing demands faced by patients.

> *“Um and then we also have a huge mental health population here, so I think part of it is we have a lot of patients that just really need a lot of help… They don’t have a lot of resources; they don’t have a lot of support. Asking them to, you know, remember to take more than one medication twice a day is asking a big thing of some of these patients. And sometimes, they just don’t have the ability to do it, unfortunately. They don’t have the skills to be taking routine medications more than once a day or even once a day. And then I think the other side to that is they just, for whatever reason, I mean, there are multiple reasons, they’re just quote-unquote done. They don’t want to do it anymore. They’re sick of taking medications. They know what this could do to their health, but they just stop. And then they come see us and we get them back on it, and they take it for another month, maybe two months, and then they fall off again.”*

#### Theme 3: Lack of familiarity (Capability)

The relative novelty of SGLT2i and GLP1a posed a barrier for some clinicians. Specifically, as clinicians are still acquiring familiarity and skill prescribing these agents, some describe feelings of uncertainty, knowledge gaps, and lack of confidence. For example, several clinicians expressed hesitation in prescribing SGLT2i and GLP1a due to uncertainty about what could go wrong with their use. When compared to metformin for adults with type 2 diabetes mellitus, which has decades of long-term data across large populations supporting its safety and efficacy, SGLT2i and GLP1a have only recently emerged, and therefore clinicians have not yet had time to accumulate their own real-world observations of the efficacy and risks associated with their use. This hesitation can also be compounded by clinicians overweighing the potential risk of side effects, such as euglycemic diabetic ketoacidosis and renal worsening with SGLT2i.

> *“…being scared of the side effects, because agents like metformin have been demonstrated over decades to be rather safe and effective. And then, when you learn about these new medications, you learn about their side effects and that kind of puts people off. Like, you’ll see one patient coming in with euglycemic [diabetic ketoacidosis] from an SGLT, and that’ll leave, kind of, a bad imprint in your mind whenever you think about the medication again.”*

Statements like this link the power of adverse events on clinicians’ memory and decision-making processes with clinicians’ beliefs about the consequences of medication use and emotional concerns about causing patient harm––all amplified by a lack of familiarity with the medications. One resident suggested that headline bias may be a contributing factor to apprehension surrounding side effects:

> *“Another thing is I think even some providers are overly concerned and fearful of the potential side effects of these medications because of how much they are magnified in the media.”*

When side effects do appear, some clinicians report being uncertain how to handle them, with one nurse practitioner observing that clinicians are quick to discontinue use of the medication, instead of returning to a lower, tolerated dose. Among interviewed clinicians, residents more commonly cited lack of practice prescribing SGLT2i and GLP1a and lack of exposure to the medications being used in different clinical contexts, such as for adults without diabetes.

Moreover, the interviews revealed two main knowledge gaps surrounding the use of SGLT2i and GLP1a: (1) determining appropriate clinical contexts and (2) selecting the most suitable agent within each class. For example, some clinicians expressed uncertainty about initiating SGLT2i therapy in patients with chronic kidney disease but no diabetes, including questions around eligibility criteria and appropriate timing. Another challenge involves weighing the need for insulin with the desire to offer the protective benefits of SGLT2i or GLP1a in patients with severely elevated A1c levels:

> *“I guess one of the barriers that we have with that is when people don’t really understand that med and we put patients on it too early in their diagnosis, because when their sugars are through the roof, I find that those patients tend to get worse infections and then they hate the med, they won’t go anywhere near it, and then when you talk to them about it a year later, when they’re well-controlled and they would benefit from it, they’re like: ‘Nope, I won’t go anywhere near that.’”*

Beyond this, some providers described choice paralysis when selecting a specific agent within each drug class, owing in part to information overload and a rapidly-expanding landscape of available diabetes medications.

Finally, a few clinicians reported feeling less confident prescribing SGLT2i and GLP1a because they are still developing the skills needed to prescribe and manage these medications. This includes understanding the different titration schedules of individual agents, overcoming roadblocks related to medication coverage, and knowing when to hold and restart treatment around hospital procedures. Altogether, lack of experience prescribing these medications seems to create a sense of greater effort when starting them for patients.

> *“The SGLT2s, I think like, you know, it’s an oral route and, you know, I’m more familiar with the dosing. Like, I think it’s something… I think it’s just more that there’s less of a mental barrier for me to prescribe those for whatever reason. I think I have less comfort with the GLP1s so far, but that’s something I’m obviously hoping to work on. I tend to have to look [GLP1a] up each time.”*

#### Theme 4: Clinical identity (Motivation)

Clinicians’ personal identities were a prominent underlying factor to some commonly cited barriers to SGLT2i and GLP1a prescribing. Personal beliefs about one’s role in the care of those with chronic conditions and personal practices (including adherence to clinical guidelines) limited prescribing in the following ways.

For one, clinicians’ individual risk tolerances factor heavily in their willingness to prescribe these agents. Although all clinicians reported encountering patient side effects, these experiences do not necessarily hinder clinicians from prescribing these agents overall; it is only for certain patient groups that their clinical judgement deems the risk of prescribing to exceed the expected benefits. In withholding a prescription, clinicians hope to avoid side effects which may cause patient harm. For example, elderly patients are one group for whom clinicians often describe reluctance to prescribe, citing concerns about cognitive decline, challenges with sick day management, and risks such as hypovolemia, unintended weight loss, and frailty.

> *“People can get pretty dehydrated from it if they’re not paying attention to that. So, you know, older patients, patients with cognitive impairment—might not be the best medication for them. I’m kind of scared that it’ll just—in the nursing home, just getting all these medications in a blister pack, their caregiver’s not going to think of holding it, then it’s probably a little bit more dangerous in that case.”*

Other patient groups for whom clinicians exercise prescribing caution are listed in **Appendix C**.

Second, some clinicians’ perceived scope of practice makes them feel like SGLT2i prescribing falls outside of their responsibility. This idea came up for a few providers when asked about managing patients with chronic kidney disease without diabetes, with clinicians seeing this as falling more within the domain of a nephrologist:

> *“Heart failure, yes. Chronic kidney disease, no. I refer renal patients to the nephrologist. We actually have pretty good access to nephrologists.”*

Similarly, the emergency physicians we interviewed described SGLT2i and GLP1a prescribing as beyond their scope, citing limited capacity for follow-up and a desire not to overstep clinical roles by initiating these medications ahead of primary care providers.

Third, personal beliefs tie into clinical practice when it comes to prescribing GLP1a as a weight loss option for adults who are overweight or obese––one area in particular where clinicians described hesitancy in accepting the use of these medications. While most clinicians acknowledged the evidence supporting GLP1a use for weight loss and observed an uptick in patients explicitly inquiring about semaglutide, their personal beliefs remained rooted in advocating for behavioral strategies, including diet and exercise, before considering a GLP1a prescription.

> *“I do have a lot of patients asking about Ozempic [GLP1a] now, unfortunately, but I don’t—like I still try to push the lifestyle and the healthy eating and all of that, but if they’re an inpatient, then I may just ask for an endocrinologist consult, but that’s if my patient is really sick and they’re unable to really like exercise, like if there’s no other options, then I’ll push towards that…”*

Some resident physicians who expressed these opinions referred to their preceptors sharing this perspective, suggesting a possible social influence. One resident further pointed out that their preceptors are reluctant to initiate GLP1a treatment for weight loss because they do not believe it to be a permanent solution, and perceived ongoing stigma in perceptions of obesity as a chronic disease that requires pharmacological management.

> *“I have seen other preceptors, not my main ones, say…almost in a very negative way that this will never help a patient. Like for example, like Victoza [GLP1a], Ozempic [GLPa], or Trulicity [GLP1a], you know, giving a patient a medication like this is not going to help obesity, because if they stop it, they’re going to go right back. And, yeah, maybe there is some truth to that, because it’s also like a psychological aspect with obesity and weight gain, but there’s other aspects too, right?”*

Fourth, in a few interviews, clinicians mentioned that the clinical guideline-directed use of metformin as first-line therapy for diabetes sometimes becomes a barrier to initiating SGLT2i and GLP1a because it makes clinicians feel like they are deviating from established recommendations that they should follow.

> *“For the SGLT2s, barrier would probably be, yeah, I think sometimes, there’s still that barrier of metformin being, you know, still the go-to first for a lot of the guidelines or just historically. So, jumping straight to that, you feel like you’re cheating a little bit as a prescriber.”*

Given the robust nature of data on SGLT2i and GLP1a’s multifold benefits, some clinicians questioned why metformin remains first-line therapy and anticipated that future guidelines may prioritize these newer therapies instead.

#### Theme 5: Behavioural inertia (Capability)

The final major barrier that emerged from the interviews was the transitional period of behaviour change needed to actually incorporate these newer therapies into routine practice. This time lag between learning about these newer medications and prescribing them suggests that more than just knowledge may be required to drive behaviour change.

> *“Yeah, I think it’s just an omission on my part, and if I think about it, because I did read the last study on it, which was actually very compelling. I guess it just hasn’t translated yet into practice for me.”*

### Facilitators

#### Theme 1: Role as a clinician that follows the data (Motivation)

Generally, clinicians reported familiarity with and common use of SGLT2i and GLP1a in their practice as second-line agents for adults with type 2 diabetes and cardiorenal risk factors or high body mass index. Most endorsed using SGLT2i as one of the four pillars of heart failure management and some described prescribing SGLT2i for individuals with chronic kidney disease. Among frequent users of these agents, many referred to current clinical practice guidelines as directing and providing clarity on prescribing decisions, including Diabetes Canada guidelines, the Canadian Cardiovascular guidelines, and Kidney Disease: Improving Global Outcomes guidelines for chronic kidney disease. Some clinicians’ statements suggested that their clinical role is one closely aligned with following these straightforward and trusted clinical recommendations.

> *“I’m very guideline based….I feel like the guidelines are just so helpful. When they come out with very clear guidelines, it just makes it so much easier, especially because— especially in primary care. At my clinic, we are dealing with so many different things, and so diabetes is just one aspect of it, and there are so many new drugs it seems like are always on the market. So the guidelines just make it more like—I feel like as NPs in general, we’re more guideline-based.”*

Similarly, clinicians who view themselves as evidence-driven shaped their prescribing behaviour off of data emerging from recent trials on the agents’ benefits.

> *“And especially like the hard endpoint data that’s being brought through these trials, so looking at reduced hospitalizations, reduced cardiac or renal failure, so I think, for those especially looking at the trial data and trying to incorporate that into your practice, but also keeping in mind the patient in front of you.”*

#### Theme 2: Belief that SGLT2i/GLP1a will improve patient outcomes (Motivation)

Two powerful facilitators of SGLT2i and GLP1a prescribing include, first, the desire to offer patients medications with strong benefit profiles, and second, optimism that SGLT2i and GLP1a use will meaningfully improve patients’ health outcomes. Most clinicians reported considering cardiovascular, renal, and body mass index risk factors in their prescribing algorithms and advocating for SGLT2i and GLP1a use on the basis of their risk reductions and protective effects.

> *“I’m way more concerned about the cardiorenal benefits. And there certainly are times I have to add something more, but it’s not to reach an A1C of 6 ‘cause that kills people. It’s more around the cardiorenal protection….I-I present the two (empagliflozin [SGLT2I] and Ozempic [GLP1A]) to the patient. I do believe [in shared decision-making] and I will tell the patient that the…empagliflozin has been shown to have a greater cardioprotective effect and a greater renal effect, so for me, it would probably be the first choice, but again, bringing in the patient’s values of worrying about bladder infections, weight loss, that would be involved in the decision.”*

This quotation highlights an important nuance of this theme, which is that while some clinicians explicitly encourage SGLT2i and GLP1a as the optimal choices for their patients, their final prescribing decision ultimately takes into account patient preferences and concerns.

Clinicians’ explicit intention to prescribe these medications were often coupled with positive perceptions of the medications’ ability to improve health outcomes. Optimism about immediate results was generally informed by experiences in which patients had A1c reductions, lost weight, and tolerated the medications well, while confidence in long-term benefits was grounded in knowledge of clinical trial data.

> *“I think the benefits are strong and strongly outweigh the risks. I’m very taken back and impressed with how the SGLT2s have shown these benefits that you never thought they would have. It’s quite remarkable how it is having a survival–– like a mortality benefit, like hard outcome differences, which are very hard to come by in medicine, especially for heart failure and CKD, which we never expected.”*

#### Theme 3: Clinicians’ perceptions of patient openness (Opportunity)

Previously, we highlighted patient refusal as being a significant barrier to starting SGLT2i and GLP1a. However, some clinicians suggested that patients can also be enablers of prescribing. When clinicians perceive patients to be willing to start on these agents, it becomes easier to prescribe. Notably, many clinicians reported an increase in patients asking to be prescribed semaglutide for the weight loss benefits. Not only can patient preference lead to a GLP1a selection when offered a choice between second-line agents for diabetes, but patient interest can facilitate earlier conversations about adding a GLP1. The media––particularly semaglutide advertisements––was identified by participants as being a powerful background influence in this case. One nuance to clarify here is that patient interest in semaglutide only enabled greater GLP1a prescribing when the clinician was already supportive of GLP1a prescribing for weight management.

> *“I guess now I do tend to go to the GLP1s because they know about them and they want them, so they’re preferred.”*

Additionally, clinicians suggested that patient expectations can influence prescribing. Specifically, two clinicians mentioned that introducing their diabetic patients early to the idea that second-line agents may be needed later in their care can increase patient readiness when further treatment is indicated.

> *“…now more than prior, [GLP1a are] probably in the conversation a little earlier on. And we at least bring it up to the patient and tell them…this is something we can get benefits from, just from a heart perspective, and then that way, if there’s any lack of control or there’s needed benefits that we can draw from it, patients are ready to start taking it.”*

#### Theme 4: Comfort prescribing (Capability)

Clinicians generally expressed confidence prescribing SGLT2i and GLP1a. The sources of their comfort with the medications were multifold. For one, clinicians described that having lots of experience prescribing these agents built their skill and confidence. Notably, some providers described that more experience prescribing specifically helped address their fears about side effects and challenges handling the medications, which were two barriers quoted by other clinicians and are noted above.

> *“Right now, at this point, I’ve met enough patients to be more confident, to know exactly how, you know, how we up the medication, how the medication is taken, what potential side effects are we dealing with, how long it takes for them to subside. And then what’s the outcome we are looking for and at what point we decide if this is worth it or not. So, all these questions get answered after working with just a few patients.”*

Another nuance to this is that some clinicians have acquired familiarity primarily with one agent in each drug class––typically, empagliflozin and semaglutide. This again mirrors a barrier: information overload of all the different medications in each drug class becomes challenging to navigate when deciding to prescribe an SGLT2i or GLP1a.

> *“But then, it kind of just, if I’m being honest, it just turns into you kind of pick one that you’re familiar with, that you’re comfortable with, that maybe you’ve seen or inherited people who are already on it and you go with it. I know, like with the SGLT2s, Jardiance [empagliflozin] would be kind of my go-to because I’m more comfortable with it, but as far as, like, the other ones, you know, I know they’re there.”*

Moreover, several clinicians indicated that the increased prescribing of SGLT2i and GLP1a by their colleagues fostered their own prescribing confidence, highlighting the influence of social norms in promoting knowledge-building, creating emotional ease, and promoting behaviour change. Participants perceived the media as playing a substantial role here in creating a sense of familiarity with the medications––namely, semaglutide (i.e., Ozempic).

> *“I think now just like with everything, obviously, with the media and the weight loss with the Ozempic [GLP1a], I tend to go more towards using the Ozempic, to-for weight loss – for obesity, for sleep apnea…those kinds of scenarios.”*

Another source of comfort arises from the simplicity of prescribing SGLT2i and the class being widely recommended within resources commonly used by physicians daily.

> *“I feel really confident with the Jardiance [SGLT2i], after seeing it on UpToDate, and just knowing that, that, it’s like renally—like, you don’t need a renal dose for it, it’s safe….and then there’s just the one dose that was suggested, the 10 mg. So I’m like, ok, can’t go wrong with this.”*

For some clinicians, confidence with prescribing is generated by having strong clinical trial data supporting their decision to prescribe these agents.

> *“For myself personally, if the GFR is above 20 or 25 for dapa and empa [SGLT2i] respectively, then I’m more than comfortable prescribing it, knowing what these clinical trials have shown as far as delaying the onset of dialysis.”*

For other clinicians, identifying that these medications fall under their responsibility to prescribe motivates comfort prescribing and provides a sense of comfort with the behaviour.

> *“…you have to become really comfortable prescribing these and I think they’re perfectly in the wheelhouse of family medicine.”*

#### Theme 5: System and Colleague Supports (Opportunity)

Although acquiring financial coverage for these medications was among the most highly cited barrier, some clinicians did mention that expanding eligibility criteria for drug coverage under the Ontario drug plan has increased opportunities for prescribing.

> *“I think the biggest thing is that they have become so much more accessible in the last few years. I remember before, you know, they first came out 5 years ago, that you could not get people on these unless you met all this criteria; you’ve tried this, this, and that. Like, glyburide and gliclazide, which nobody should be on, right? It made care very onerous, and I think it’s overall positive that the, you know, eligibility has been expanded to keep up with best practice and best evidence.”*

Learning about SGLT2i and GLP1a prescribing through colleagues also proved to be an important influence motivating clinicians to incorporate these medications into routine practice. Residents described adopting the prescribing behaviors of their preceptors, gaining familiarity with these medications in the clinical contexts to which they are exposed. For many of the interviewed clinicians who were taught only about metformin, insulin, and sulfonylureas while in school, all of their learning about new medications happens on the job, with colleagues (including colleagues in their clinic, internal medicine colleagues, and specialists like cardiologists and nephrologists) playing a large role as social enablers of prescribing.

> *“…and then a lot of times as well, I’m gaining a lot of information from consult notes that I get back from nephrology or cardiology, saying: ‘hey let’s do x, y, z because of a, b, c.’ Which is often: ‘I’ve added this on because of CKD’ or ‘I’ve added this on because of high risk of cardiovascular disease.’ This is sort of how the learning goes on…”*

## DISCUSSION

In this qualitative descriptive study of clinicians practicing in Northern Ontario, we identified five main barriers and five main facilitators to SGLT2i and GLP1a use for adults with chronic disease, including type 2 diabetes, heart failure, obesity, and chronic kidney disease. Among the barriers, limited access to the medications reflects system-level challenges; patient challenges and competing demands represent patient-level obstacles; while lack of familiarity, clinical identity, and prescribing inertia are provider-level concerns. The same stratifications were found among facilitators: at the system-level, expanding coverage and multi-disciplinary collaboration are key; at the patient-level, patient interest and readiness encouraged prescribing; and at the provider-level, a data-oriented clinical identity, optimism about SGLT2i and GLP1a improving patient outcomes, and confidence prescribing were elements driving the prescription of these medications. Taken together, these results highlight multiple opportunities for knowledge translation activities with an emphasis on education that could reduce modifiable barriers and amplify existing facilitators of prescribing.

### Contextualizing our results within the broader literature

In 2021, a qualitative semi-structured descriptive study conducted in Australia identified that family physicians lacked familiarity with SGLT2i and preferred their endocrinologist colleagues to initiate these medications for their patients with diabetes.^38^ At the time of our study, conducted three years later, knowledge and widespread prescribing of these agents appears to have improved. All interviewed clinicians were aware of the medications’ cardiorenal benefits, with remaining knowledge gaps pertaining mainly to specific patient populations––such as those with chronic kidney disease but without diabetes. However, our study showed that two of the barriers identified in 2021 persist today: both the rapidly evolving drug landscape for diabetes management and patients’ experiences with side effects continue to be barriers (the latter primarily causing patient intolerance to the medication rather than clinician reluctance to prescribe). Moreover, population-based cohort studies indicate that there is generally lower use of these agents among older adults, women, and those with lower household income.^28,39^ Our study suggests that some of the reasons for lower prescribing rates among these groups include clinicians’ wishes to avoid harms (i.e., side effects), patient fear of urogenital infections with SGLT2i use, medication cost, and low medication adherence.

### Clinician knowledge and beliefs as strong facilitators to use of SGLT2i and GLP1a

One facilitator strongly emphasized by several clinicians was their belief that SGLT2i and GLP1a are effective medications that can improve the health outcomes of their patients by reducing their risk of adverse cardiovascular events, progression to end stage renal disease, and mortality. When directly asked about barriers to SGLT2i and GLP1a prescribing, many of these clinicians listed experiences with patient side effects (more commonly with GLP1a use, less commonly with SGLT2i use). However, many clinicians were adamant that these experiences do not hinder them from prescribing these agents, since they believe the benefits to largely outweigh the risks. Thus, clinician optimism about the medications’ protective effects is a powerful motivator of prescribing, and should be appealed to in any knowledge translation activities aimed at improving SGLT2i and GLP1a uptake.

Knowledge accrual was a key contextual enabler of SGLT2i and GLP1a prescribing. Our interviews revealed several different pathways through which clinicians acquire and update their knowledge on SGLT2i and GLP1a, including both formal and informal channels. Knowledge of the medications was found to underlie most clinicians’ comfort prescribing these agents by normalizing their use in best medical practice and informing their beliefs that these agents will improve patient outcomes.

### Limitations to access in Northern Ontario as a strong barrier to use of SGLT2i and GLP1a

Limited access to SGLT2i and GLP1a emerged as the most significant barrier to prescribing in Northern Ontario. This challenge reflects broader systemic issues, including the high proportion of patients in Ontario who lack a primary care provider and thus have no regular point of contact with clinicians who could initiate these therapies. Among those who do access care, patient eligibility for drug coverage weighed particularly heavily in providers’ prescribing algorithms. Two groups identified as being disproportionately affected by insufficient drug coverage included patients who were unemployed or of low socioeconomic status and those seeking semaglutide for weight loss without a diabetes diagnosis. In both cases, individuals stand to benefit substantially from treatment.^1,2^ These findings therefore underscore structural barriers that exacerbate health inequities, particularly in rural and remote settings, and suggest that improving both access to primary care and equitable drug coverage would meaningfully expand the reach of these medications. Prior studies have shown that medication cost, in particular, is a major barrier to accessing GLP1a and SGLT2i.^40^

### Reluctance to prescribe GLP1a despite their benefits

Our study identified that there is lingering resistance among some clinicians to prescribe GLP1a for weight loss. This reluctance to prescribe may be influenced by the exclusion of many of these patients from cost-covering programs and may be partly explained by the persistent stigma surrounding obesity. Medical education and the media often frame obesity as a matter of personal responsibility, emphasizing lifestyle interventions such as diet and exercise, while giving less attention to the metabolic dysregulation underlying this disease and the proven efficacy of pharmacological therapies in reducing long-term health risks. This overemphasis on willpower-driven behavioral change further contributes to inequities in the provision of weight loss pharmacotherapies that have long been documented.^41,42^ Dismantling some of this stigma through education should be a crucial tenet to increasing patient access to GLP1a.

### Informing future knowledge translation activities

Drawing attention to facilitators and barriers that directly mirror one another may also guide the development of knowledge translation activities. For example, at the patient-level, clinicians noted that patients who refuse SGLT2i or GLP1a out of a desire not to increase their pill burden present a clear barrier to prescribing. However, a suggested facilitator was introducing patients early in their care to the possibility of needing a second-line agent and counselling on their cardiorenal benefits, which may increase patient willingness to start these therapies if later indicated.

At the provider-level, clinicians’ perceived scope-of-practice boundaries sometimes shifted prescribing responsibility to other clinicians; however, in the context of provider shortages––particularly family clinicians––this may leave patients with no one to initiate therapy. Thus, expanding education on the potential role of emergency physicians in Northern Ontario in managing diabetes for those without primary care providers, or of family physicians in chronic kidney disease management, could help bridge these missed opportunities to prescribing SGLT2i and GLP1a for patients who stand to benefit.

Additionally, experience with the medications created a continuum of confidence prescribing them: insufficient familiarity prescribing breeds fears and uncertainty, whereas on the other end of the continuum, greater experience prescribing facilitates behaviour change, increases confidence managing these medications, and reduces concerns about side effects. Collectively, these findings suggest that early patient engagement, targeted education, and opportunities to gain prescribing experience could increase prescribing.

Interestingly, diabetes education centres were not explicitly identified as facilitators of SGLT2i and GLP1a prescribing. While clinicians spoke positively of these centres’ support with insulin teaching, insulin management, and patient follow-up, they did not describe these centres as promoting or facilitating the use of SGLT2i or GLP1. We acknowledge that this may not reflect the experience of all providers in Northern Ontario and that diabetes education centres not represented in our interviews may actively support SGLT2i and GLP1a prescribing. One family physician even proposed that diabetes educators in Northern Ontario should have greater prescribing authority to reduce the burden of diabetes management on primary care providers.

### Study strengths and limitations

Our study has multiple limitations. First, we did not capture and code video footage of the interviews, so it is possible that we missed important emotional undertones in clinicians’ responses. Second, this study reflects the experiences of a small sample of clinicians working in Northern Ontario, and thus findings may not apply to urban centres where clinicians may face unique facilitators and barriers to SGLT2i and GLP1a prescribing. Third, we lacked the patient perspective directly. However, we believe the provider experience was sufficient to capture numerous patient-level challenges with SGLT2i and GLP1a uptake, informed by real patient-provider conversations regarding next steps in care. One strength of our study was our ability to capture the experiences of clinicians working in numerous cities across Northern Ontario and in various clinical contexts, including family medicine clinics, nurse practitioner-led clinics, diabetes education centres, hospitals, and emergency departments. We hope that this improves the generalizability of our findings.

### Summary of findings through the COM-B model and Conclusion

The insights gathered through these interviews suggest that clinician motivation and capability are strong drivers of SGLT2i and GLP1a prescribing. Clinicians who refer to clinical trial evidence and guidelines and those who learn nuances about prescribing SGLT2i and GLP1a through their colleagues were generally more optimistic about these medications, more comfortable prescribing them, and stronger advocates for their use with their patients. Even so, it is interesting to note that the barriers identified in our interviews persist despite the generally strong knowledge clinicians have about SGLT2i and GLP1a. This highlights that the main barriers to prescribing primarily relate to opportunity (e.g., financial barriers, drug shortages, patient preference) and personal factors (e.g., individual risk tolerance, personal acceptance of these medications).

Thus, the creation of knowledge translation tools should focus on increasing the accessibility of summarized information on SGLT2i and GLP1a to improve patient education and reinforce provider confidence when prescribing these medications in diverse clinical scenarios. Importantly, greater dissemination of information aimed at dispelling stigma related to obesity could facilitate GLP1a prescribing for many who stand to benefit from the medication’s multifold protective benefits. While overcoming system-level barriers such as drug coverage and clinician shortages are much greater challenges requiring policy-level interventions, knowledge-building on the safe use of SGLT2i and GLP1a can address patient- and provider-level concerns and increase the uptake of these highly beneficial therapies for adults both with and without diabetes.

## Data Availability

An aggregate summary of the data generated during this study is presented in this published manuscript. Individual data transcripts cannot be publicly shared due to confidentiality.

