## Appendix for "Facilitators and barriers to SGLT2i and GLP1a prescribing in Northern Ontario: a qualitative interview study"

**Appendix A: Semistructured Interview Guide**

Hello, I am [*introduce self and role*]. It's nice to meet you. Before we get started, I would like to take this opportunity to remind you about some important points regarding today's interview.

First: Our study aims to look at the uptake of the newer diabetes medications SGLT2 inhibitors and GLP1 analogues in Northern Ontario. These interviews are intended to help us get some insight into the circumstances and challenges you have been facing in prescribing these medications when caring for adults with and without type 2 diabetes mellitus.

Second: Your participation in this study is entirely voluntary. You are free to skip any question you do not wish to answer and if at any time during the interview you wish to stop participating, you are free to do so.

Third: The interview today will be audio recorded. I will ask that you please turn off your camera and we will only begin audio-recording after you provide informed verbal consent to participate. We ask that you do not give your name or the names of any patients or colleagues over the course of the interview. After the interview has completed, your audio files will be de-identified, transcribed, and then deleted.

Finally: Although your identifying information will be removed from all data, we cannot guarantee absolute confidentiality of information.

Do you have any questions for me at this time?
We will now move to verbal consent:

At the bottom of the information letter attached to our email correspondence, there is a verbal informed consent script. Could you please confirm that you have read this and that you provide verbal consent to participate in our study?

Yes/No

*Note: This is a semi-structured guide as follow-up questions or prompts may be used through the course of the discussion.*

**[0] Which of the following categorizations applies to you?**

(a) Physician;
(b) Resident physician;
(c) Nurse practitioner.

**[1] For a patient with T2DM requiring medication, how do you decide what to choose?**

- Prompt: What knowledge/recommendations/guidelines/evidence do you draw on when prescribing medications for patients with T2DM? Why do you choose to refer to this/these resource(s)?
- Prompt: Do you ever refer to your colleagues' advice?
- Prompt: How do you prioritize different resources (e.g., colleagues vs guidelines)?

**[2] For a patient with T2DM who is already on a medication for their diabetes, how do you decide when to add another medication?**

**[3] If the patient is overweight or obese, how does that change your management?**

**[4] How does your management differ for a patient with T2DM who also has coronary artery disease, heart failure, renal failure, or stroke?**

- <u>If they don't say SGLT2 or GLP1</u>
    - Are you familiar with SGLT2 or GLP1?
- <u>If they are familiar with them:</u>
    - There are so many new classes of drugs, how did you hear about these drugs? (Guidelines? Studies? Colleagues? Other?)
    - What can you tell me about these drugs?
    - How confident are you prescribing them?
    - In your experience, are these classes of medication effective?
    - Have you observed any incentives to start these medications?

**[5] When considering a newer agent like an SGLT2i or GLP1a, what do you see as the biggest challenges or barriers to their use?**

- <u>If they don't use them/aren't familiar with them</u>:
    - Are there particular barriers you have identified?
    - Do your colleagues prescribe them?
    - What would change your practice to prescribe these medications?
- <u>If they use them:</u>
    - How did you acquire familiarity with these classes of medications? (Guidelines? Studies? Colleagues? Other?)
    - Do your colleagues prescribe them?
    - In your experience, are these classes of medications effective?
- Are there certain side effects that you worry about? Do these concerns lead to you not prescribing SGLT2i? GLP1a?
- Are these medications reasonable for a family doctor or nurse practitioner to prescribe, or is the expectation that an endocrinologist will?
- Is it encouraged by family medicine organizations to prescribe these drugs?

- Is cost a barrier?

**[6] Have you prescribed either class of medications for adults without diabetes?**

- If yes, tell us more.
- If not, were you aware that SGLT2i are effective for heart failure and CKD regardless of T2DM? GLP1a for adults with obesity regardless of diabetes?

**[7] For your average patient with T2DM who you start on an SGLT2i, what do you think their risk of DKA is?**

- If they don't know, prompt them with 1 in 50? 1 in 70? 1 in 100? 1 in 1000? 1 in 10,000?

**Concluding the Interview**

- Summarize some key points from the interviewee's responses
- Invite interviewee to share some final thoughts
  - Prompt: Is there anything else you would like to share about your experience prescribing medications for adults with and without T2DM that we haven't already touched upon?
- Do you have any questions before we conclude the interview?
- Thank the interviewee for their time.
  - We would like to offer you a **gift card honorarium** to thank you for your time today. With your permission, we will send this gift card electronically to your email immediately following the conclusion of this Zoom call.
- Cover next steps (transcribe the interview, delete any audio recordings, if interested in receiving results, please reach out to the research team)

**Snowball Sampling**

- If you know of anyone who may be interested in participating in this interview study, we would appreciate it if you could please pass our recruitment email forward.

### Appendix B. Codebook.

| Parent Code and Sub-categories | | Definition [a] | Example Quote |
|---|---|---|---|
| **1. Knowledge** | | **Knowledge or awareness of SGLT2i and GLP1a.** | |
| | 1.1. Knowledge of practice guidelines. | Knowledge of practice guidelines (e.g., Diabetes Canada, medical school) that currently guide prescribing or may be referred to for guidance prescribing SGLT2i or GLP1a. | *"As far as guidelines, [Canadian Cardiovascular Society] for cardiac stuff, [Kidney Disease: Improving Global Outcomes] for kidney, and Diabetes Canada has excellent guidelines as well as an application that you can download on your phone and I do use that as well."* |
| | 1.2. Knowledge of evidence base/cardiorenal benefit. | Knowledge of the key trials on SGLT2i and GLP1a or knowledge of the benefits associated with SGLT2i and GLP1a use in adults with and without diabetes arising from these key trials (e.g., EMPA-REG, EMPA-Kidney, CREDENCE, DAPA-CKD, DELIVER, FLOW, SELECT). This knowledge may have been acquired directly from published literature or via podcasts, conferences, etc. | *"I'm a lot more well-versed with the cardio trials, but for SGLT2i, I know EMPORER-reduced, DELIVER, DAPA-HF are the big ones for heart failure, and for SGLT2s in kidney disease, there's EMPA-Kidney. I think that's the main one I refer to."* |
| | 1.3. Knowledge of mechanism of action. | Knowledge of the physiologic mechanisms of action of SGLT2i and GLP1a, which may inform what clinicians expect of the medications' benefits and side effects. | *"…how I explain it is that, it works directly in your kidneys to…to ensure that sodium and specifically glucose—so salt and glucose—is excreted more….And I say that to the patient to let them know that because this is happening, the side effects of having more yeast infections are there. Also, you want to remain hydrated because of, you know, losing more sodium."* |
| | 1.4. Knowledge of clinical indications. | Knowledge of the clinical indications for SGLT2i and GLP1a, including type 2 diabetes, heart failure (regardless of ejection fraction), chronic kidney disease, and obesity, as well as the medications' contraindications. | *"…if they're not at their A1C target, and then also if…yeah, if they have some kind of, if they have coronary artery disease or they have, like, CHF… or, you know, they have obesity and they'd like to lose weight…or they have…they're progressing toward CKD, I might add these medications in. Even if they are at their A1C target, I think it would be reasonable to add these medications."* |
| | 1.5. Knowledge of using the medication | Knowledge of handling SGLT2i and GLP1a (e.g., agent selection, dosing, titration, holding the medication before procedures, etc) for their patients. | *"I never know when to start titrating off something else."* |

| | | | |
|---|---|---|---|
| **2. Skills.** | | **Competence in counselling on, prescribing, and handling SGLT2i and GLP1a for those who may benefit.** | |
| | 2.1. Familiarity. | The clinician describes skill prescribing SGLT2i and GLP1a. Skill may have been acquired over time and experience prescribing. This category includes uncertainty prescribing SGLT2i or GLP1a in certain contexts because of limited experience in such contexts (e.g., prescribing for patients with CKD). | *"Right now, at this point, I've met enough patients to be more confident, to know exactly how, you know, how we up the medication, how the medication is taken, what potential side effects are we dealing with, how long it takes for them to subside. And then what's the outcome we are looking for and at what point we decide if this is worth it or not. So, all these questions get answered after working with just a few patients."* |
| | 2.2. Handling the medication. | The clinician describes comfort and skill dosing, titrating, managing drug-drug interactions, holding medications when required, and arranging appropriate follow-up for SGLT2i and GLP1a, based on patient characteristics (e.g., kidney function, polypharmacy, age, etc). | *"I tend to ask a lot of questions about how quickly I can change dosing on things. When I worked purely outpatient, we would see patients every 3 months, so it was kind of easy. You would just increase the dose next time you saw them. You wouldn't be increasing as frequently as you can when you work in a hospital setting."* |
| **3. Social/professional role and identity.** | | **Personal qualities, beliefs, or behaviours attached to one's professional identity and/or professional role.** | |
| | 3.1. Role in diabetes care. | The clinician describes their personal approach to managing those with type 2 diabetes. Their professional role may include achieving glycemic control and/or protecting against cardiorenal complications. | *"-- so part of me was, part of my job was to kind of de-prescribe these and switch people over to things that offer heart protection as well as kidney protection."* |
| | 3.2. Role in caring for those with heart failure, chronic kidney disease, and/or obesity. | The clinician describes their personal approach to managing those with heart failure (regardless of ejection fraction), chronic kidney disease, and/or obesity. | *"...the standard teaching is to start with metformin. I think in my experience, I'm always in an inpatient setting and what I would say is I'm much more inclined to be treating the comorbidities, namely the cardiac and CKD."* |
| | 3.3. Role in referring to a primary care provider or specialist for prescription of an SGLT2i or GLP1a. | The clinician describes that prescribing SGLT2i or GLP1a for adults with or without diabetes falls outside their clinical responsibility, with their role being to refer patients to a primary care provider or specialist for a prescription. | *"...if they're complex medical patients, we are typically sending them to our medical clinic, and then again, they need some follow up with these medications and I don't have the capacity to do that."* |
| | 3.4. Role as a clinician. | The clinician describes their alignment to professional expectations or values, including but not limited to patient-centred care, informed decision-making, evidence-based care, and respecting clinical guidelines. | *"We have to give the evidence to people."*<br><br>*"...making sure that we use medications that are renally [protective] as well as offer cardioprotection."* |

| | | |
|---|---|---|
| 3.5. Identity as a clinician. | Participants use more personal language relating to their perceived role and identity as a clinician. This may include language relating to "who I am" and "what being a clinician is all about." | *"I think that I'm definitely an advocate--just based on my background. I've done Kinesiology as my undergrad and I'm kind of aware of some of the research around exercise, physical activity....More and more, I think cardiorespiratory fitness and weight are risk factors that I think we need to address a little bit more aggressively than we have been."*<br><br>*"I'm very guideline based –"* |
| **4. Beliefs about capabilities.** | **Self-confidence and perceived competence or ability.** | |
| 4.1. Confidence in prescribing | The clinician is confident counselling on and prescribing SGLT2i and GLP1a for adults or describes experiences/outcomes that have reinforced or changed his/her confidence in prescribing. | *"Right now, at this point, I've met enough patients to be more confident, to know exactly how, you know, how we up the medication, how the medication is taken, what potential side effects are we dealing with, how long it takes for them to subside. And then what's the outcome we are looking for and at what point we decide if this is worth it or not. So, all these questions get answered after working with just a few patients."* |
| **5. Optimism** | **Belief that SGLT2i and GLP1a use will attain clinical goals.** | |
| 5.1. Optimism that medication use will result in better health outcomes for patients. | The clinician believes that use of SGLT2i and GLP1a will improve long-term health outcomes for their patients, including but not limited to glycemic control, cardiorenal protection, weight loss, reduced mortality, and/or improved quality of life. | *"I would say the lack of significant side effects for most people is a big positive knowing that they can tolerate these medications and they'll have those health benefits from it. It's reassuring to me to know that we're not putting these people through a ton of intolerable side effects to achieve that…"* |
| 5.2. Good outcomes | The clinician describes experience with good outcomes arising in patients started on SGLT2i or GLP1a—including but not limited to reductions in HbA1c, improvements in blood pressure, improvements in renal function, and patient satisfaction—which reinforces ongoing SGLT2i and GLP1a prescribing. | *" …for the most part, I would have, you know, positive things to say when talking about what I know generally about these medications-- at least from the patient experience and from the data on the health outcomes....knowing that there's that benefit in place and that they're on these medications and adhering to them and not experiencing significant side effects is good…"* |
| **6. Beliefs about Consequences** | **Beliefs about outcomes and/or potential consequences of SGLT2i and GLP1a use in patients.** | |

| | | | |
|---|---|---|---|
| | 6.1. Poor or adverse outcomes. | The clinician anticipates poor or adverse outcomes for patients using SGLT2i or GLP1a (e.g., subtherapeutic effect or complications such as urogenital infections, euglycemic diabetic ketoacidosis, nausea, diarrhea, etc). This includes anticipating poor or adverse outcomes in specific patient populations (e.g., the elderly, women with frequent UTIs). These beliefs about consequences may be informed by past experiences but influence current decision-making on prescribing SGLT2i or GLP1a. | *"I think like especially with euglycemic DKA, I think that's something that's quite scary for me, as a clinician. So, if a patient seems like they're not someone that'll seek medical attention if they're unwell, then like that's something that would worry me about prescribing one of these medications. 'Cause euglycemic DKA tends to go more under the radar. Like if they check their sugars, they're not really going to notice from their sugars that they are in DKA. So like maybe a patient that doesn't seem the most responsible, that would give me a bit of hesitancy [prescribing]."* |
| | 6.2. Patient dissatisfaction. | The clinician has experience with or anticipates patient dissatisfaction with use of an SGLT2i or GLP1a, because of side effects, route of administration, etc. | *"And I've also seen mycotic infections as well. Like, UTIs and genital infections. So that's something that patients get kind of annoyed with, as well."*<br><br>*"I usually don't bring it up in the earlier stages based on my assumption, I suppose, that people generally don't like injecting themselves."* |
| **7. Reinforcement** | | **Rewards, incentives, or punishments influencing prescribing.** | |
| | 7.1. External reinforcement. | The clinician considers external incentives or reinforcement when considering whether or not to prescribe SGLT2i and GLP1a, such as continued education credits or billing. | No applicable quotes. |
| | 7.2. Policy/infrastructure | The clinician describes that their prescribing behaviour is at least in part influenced by performance incentives in the hospital or clinic in which they work. | No applicable quotes. |
| **8. Intention** | | **The conscious decision to prescribe or not prescribe an SGLT2i/GLP1a for a given reason(s).** | |
| | 8.1. Appropriate counselling | The clinician describes having thoughtful discussions with patients about their medication options for glycemic control, including SGLT2i and/or GLP1a (even though patients may ultimately refuse the medication afterwards). | *"I would try to, you know, have the conversation with them: the mortality benefits that's been shown, the side effects, that we recommend it, and then we'd start them on it."*<br><br>*"...with shared decision-making, I would discuss options, like SGLT2s, GLP1a, DP- DPP4s…"* |

| | | | |
|---|---|---|---|
| | 8.2. Improving patient outcomes. | The clinician describes wanting to prescribe SGLT2i and/or GLP1a to improve patients' cardiovascular and renal outcomes, mortality, or quality of life. | *"…making sure that we use medications that are renally [protective] as well as offer cardioprotection."* |
| | 8.3. Avoiding harm. | The clinician describes not prescribing SGLT2i or GLP1a to avoid expected harms to patients. | *"For SGLT2 inhibitors, if an individual is elderly with mobility and/or cognitive issues, I am finding that because I am and… and perhaps their glycemic control is not really optimal just yet so they're already experiencing some glucosuria and frequency because of that, I would probably avoid an SGLT2 inhibitor in that scenario."* |
| **9. Goals** | | **Articulation of goals or action-planning related to prescribing behaviors.** | |
| | 9.1. Articulates goals. | The clinician describes wanting to change something about their prescribing practice. They may outline goal-directed plans or actions regarding their prescribing behaviour. Examples include wanting to learn more about SGLT2i or GLP1a or counselling about SGLT2i or GLP1a more broadly among patients. | *"I think I have less comfort with the GLP1s so far, but that's something I'm obviously hoping to work on."* |
| **10. Memory, attention, and decision processes.** | | **The role of one's prior experiences, attention in prescribing, and ability to select between treatment options.** | |
| | 10.1. Memory and attention in prescribing. | The clinician describes consolidating clinical experiences and knowledge of guidelines or clinical trial data to make informed decisions. | *"…if someone has an A1C of 12, I'm not going to start empagliflozin, because I have done that in the past and it seemed that everyone was getting yeast infections. And then, they're against starting it ever again for the rest of their life if that happens. So, I usually try to get their A1C somewhat closer to something reasonable before I start it. Otherwise, I'll turn them off the med forever."* |
| | 10.2. Patient involvement in decision-making. | The clinician shows awareness that decision-making processes regarding medication use rely on patient involvement. | *"…in choosing between them, I do involve the patient in the decision-making process."* |
| | 10.3. Risk-benefit analysis. | The clinician articulates some risk-benefit analysis when describing their decision-making processes regarding the prescription of SGLT2i and GLP1a. | *"So Jardiance would be the next one…if they're---unless, you know, they're having tons of UTIs."* |
| | 10.4. Algorithmic approach to prescribing. | The clinician describes an algorithmic approach to prescribing, which may be based on clinical guidelines, | *"If they're younger, I'll be more aggressive. If they're older, I'll be less aggressive. And then my second add-on, usually empagliflozin is my go-to….Assuming they have coverage."* |

| | | | |
|---|---|---|---|
| | | clinical trial data, and/or previous experiences using the medications. | |
| **11. Environmental context and resources.** | | **Environmental circumstances and non-personal factors that affect SGLT2i and GLP1a prescribing.** | |
| | 11.1. Connection to colleagues. | The clinician describes an ability to rely on colleagues for support prescribing SLGT2i and GLP1a, including such things as patient referrals or consultations for guidance prescribing. | |
| | 11.1.1. Relationship with nurse practitioners, residents, or other clinicians. | The clinician describes referring patients to, consulting, or sharing prescribing responsibility with colleagues, including nurse practitioners, residents and/or other clinicians, for support prescribing SGLT2i and GLP1a. | *"I can count on one hand the amount of times I've done it, because usually by that point, the specialist…people with CHF are going to have a cardiologist that's gonna [prescribe an SGLT2i]"* |
| | 11.1.2. Relationship with diabetes education centres. | The clinician describes a relationship with diabetes education centres that supports or restricts SGLT2i/GLP1a prescribing for patients with diabetes. | *"I send everyone with diabetes down to the diabetes educators for injection teaching. So I write a prescription, and they have to pick it up and bring it to the educators to learn how to use it."* |
| | 11.2. Supply. | Prescription of SGLT2i or GLP1a is limited by the availability of these drugs. | |
| | 11.2.1. Drug shortage. | The clinician describes not being able to prescribe an SGLT2i or GLP1a because these agents have been absent from local pharmacies over a certain period of time. | *"…one: availability, I guess, where I am. I couldn't believe [GLP1a] wasn't available here for months. For months."* |
| | 11.2.2. Compassionate Drug Programs. | The clinician describes that medication supply with compassionate drug programs are inadequate, which limits clinicians' ability to rely on these programs for their patients and thus, their ability to prescribe SGLT2i and GLP1a for patients who require them. | *"I don't find the Jardiance Compassionate Program very effective…. We have to fill out lots of forms, and they literally send you one month at a time… it runs out before they send you the higher dose. Right, 'cause you've got to take the lower dose for one month, then go up. And they're behind the times on sending it…. And it takes them 2 or 3 months to send the first month."* |
| | 11.3. Care setting. | Advantages or limitations inherent to the setting in which care is provided to patients with type 2 diabetes, heart failure, obesity, and/or chronic kidney disease that make it easy or challenging to prescribe a SGLT2i or GLP1a. | |

| | | | | |
|---|---|---|---|---|
| | | 11.3.1. Inpatient setting | The clinician describes a feature of working in an inpatient setting that affects their ability to prescribe a SGLT2i or a GLP1a for a patient who requires a prescription. | *"I would say that a barrier specific to the SGLT2s in the inpatient setting is that oftentimes, patients are either volume depleted or they're volume overloaded. Their kidney function is dynamic and reduced, and then goes back to baseline just at the end of their hospital stay, so when you're in this kind of clinical setting, you're a little bit more hesitant to initiate something like an SGLT2i whereas if you're seeing a patient in clinic, and they're walking in to see you and they're doing great, you're a lot more comfortable prescribing this in that setting, because their hemodynamics are much more stable at that time."*<br><br>*"I don't think I've ever actually started a patient on a GLP1a in the hospital. It's always been recommended to the family physician on follow-up because it's kind of difficult to titrate well in the hospital."* |
| | | 11.3.2. Clinician shortage | The clinician describes that SGLT2i and GLP1a prescribing at large is in part influenced by the limited availability of clinicians, including both primary care clinicians and specialists (e.g., endocrinologists, cardiologists, internists) who may be able to initiate patients on these agents. | *"..because there are so few family doctors in Northern Ontario or even endocrinologists or anyone who can help manage those things or monitor the diabetes...that some people aren't optimized because they don't have primary care to monitor or even suggest, like some people could have been like, I don't even know…"I'm on metformin and I have been but I lost my family doctor 10 years ago" and so they may not be optimized. People come into emerg all the time without a family doctor and they're admitted to hospital, don't have a family doctor, and while they're in hospital, that's when we try our best as an inpatient service to optimize their diabetes medications or get them the proper referrals to an internal medicine clinic or an endocrinologist so that they can be optimized. But I would say, a good chunk aren't because of the lack of primary care."* |
| | 11.4. Cost. | | The clinician identifies cost as a factor that restricts SGLT2i or GLP1a prescribing for patients. This includes being unable to write a prescription for those who cannot afford to pay for their medications out of pocket but do not meet Limited Use code criteria or lack sufficient drug insurance. | *"There are certain private insurances that if you don't use metformin first, they won't cover them, and they need—they have to not tolerate metformin for them to cover it."*<br><br>*"...if it's not covered by insurance, a lot of people are not able to afford it."* |

| | | |
|---|---|---|
| **12. Social influences** | **Interpersonal experiences that affect prescribing behaviour, including social pressures, norms, support, modelling, and/or interactions with others.** | |
| 12.1. Influence from a preceptor | The clinician has developed prescribing patterns or behaviours for SGLT2i or GLP1a based on what they are seeing done by their preceptor(s). | *"I will also go to my staff, like attending, for their expertise...my fastest resource is to just go to my staff and see which one they would recommend over another if I'm stuck between two."* |
| 12.2. Influence from colleagues | The clinician has developed prescribing patterns or behaviours for SGLT2i or GLP1a based on comparison or communication with colleagues (i.e., discussing indications or correct dosing of these agents.) | *"I think just the face-to-face interactions that I would typically have with internal medicine primarily in the hospital setting. I usually work in the emergency department setting, so it's a smaller portion of what I would usually do, but there's also times where I work in primary care so I think the interactions with specialists that I would have in the hospital setting often carry into the practice setting that I would have, just in clinic, so that's some of the source of information that I have."* |
| 12.3. Stigma | The clinician perceives stigma on the use of SGLT2i or GLP1a, which may be perpetuated by the media or other persons, as (1) influencing his/her own SGLT2i and GLP1a prescribing, or (2) influencing the amount of prescribing of SGLT2i and GLP1a he/she sees being done around them. | *"I have seen other preceptors, not my main ones, say, you know, like, you know, almost in a very negative way that this will never help a patient. Like for example, like Victoza, Ozempic, or Trulicity, um, you know, giving a patient a medication like this is not going to help obesity, because if they stop it, they're going to go right back."* |
| 12.4. Influence from patients. | The clinician describes the concerns, preferences, and reasons for accepting or rejecting a SGLT2i/GLP1a prescription that they hear from patients through clinical encounters. | |
| 12.4.1. Medication-averse. | These patients may be averse to polypharmacy (i.e., pill burden) or using an injection medication. These concerns may prevent clinicians from writing a prescription despite judging an SGLT2i or a GLP1a to be medically appropriate. | *"Usually, patients don't want to be on more meds…"*<br><br>*"The GLPs, you know, injections aren't people's favourite…"* |
| 12.4.2. Fear of side effects. | These patients refuse medication because of the risk of side effects or experience with side effects. | *"…I have a number of women who when you tell them it might increase the risk of yeast infection, they want to run away from it."* |
| 12.4.3. Desired health outcomes/patient preference. | These patients express specific desired health outcomes at appointments, which influences the clinician's prescribing decisions. | *"I have prescribed GLP1s for patients who don't have diabetes and primarily that's been for weight loss for patients that have approached specifically for that, saying: 'Hey, you know, I've tried x, y, z and I* |

| | | |
|---|---|---|
| | | *wanna try this and I read into the medication' and we've had a discussion about alternatives."* |
| 12.4.4. Adherence | The clinician refers to a patient's adherence behaviour or record of showing up for appointments as factors that are important to their ability to prescribe SGLT2i and GLP1a. | *"We have a lot of issues with compliance. So, I would say that's the biggest problem. If you ever end up going to North Ontario, and you go to the nursing stations, you will see fridges and fridges and boxes and boxes of, I mean, Ozempic, in the fridge that's not used, insulin that's not used. So, the biggest thing is really medication non-compliance. So I would say, we struggle, because you'll see lots of A1Cs that are quite high because a lot of people aren't using those medications...there's social things, right? They'll say "oh, they never call me 'cause of so-and-so." Like, it's actually quite nuanced, I think. There's a lot of mistrust, right? Of like the healthcare system."* |
| **13. Emotion** | **Emotion attached to prescribing choices or behaviour.** | |
| 13.1. Concern. | The clinician describes fear, worry, or concern about putting patients on an SGLT2i or a GLP1a. Language used includes an emotional component. | *"Empagliflozin I- I generally prefer, I get nervous putting women on it because they all get bladder infections and yeast infections."* |
| 13.2. Assurance. | The clinician describes reassurance or excitement about putting patients on an SGLT2i or a GLP1a. Language used includes an emotional component. | *"I felt really reassured, you know, that this is the right way to go in heart failure."* |
| **14. Behavioural regulation** | **Routine prescribing behaviours and ability to self-regulate based on new clinical recommendations** | |
| 14.1. Inertia/Routine | The clinician describes that his/her approach to managing patients or prescribing medications is based on routine (i.e., they are used to doing things a certain way or describe that "this is the way I usually behave.") | *"…before taking this interview, I was thinking about that and it- it is metformin still, despite what I just said. Probably need to ask myself why I do that."*<br><br>*"That's my personal practice and maybe something that I should change…"* |
| 14.2. Awareness | The clinician shows awareness that his/her beliefs or prescribing practice may not be aligned with the latest practice guidelines or clinical trial data. | *"I tend to be a little bit more reluctant–maybe more reluctant than I should be–to prescribe in those situations."* |

| | | | |
|---|---|---|---|
| | 14.3. Adaptation. | The clinician explicitly mentions changing their prescribing practice to include SGLT2i and GLP1a or mentions noticing clinicians around them being more conscious of opportunities to prescribe SGLT2i or GLP1a. | *"I've definitely seen, like, we're thinking about SGLT2s more and even just within a few years of me doing, like, rotations in inpatient medicine, I find, like, during rounds, if we have a patient with diabetes especially and HFrEF, then everyone's like trying to see, "oh, are they on an SGLT2?" and we're making sure that we're covering that pillar of heart failure management. So I think it's becoming more talked about and more common that we're prescribing them for that and, like, double checking that they're on them."*<br><br>*"...now it's been pretty ingrained to do the secondary benefits, like the cardio-renal benefits, so I do try to have most of the patients, I look at cardiovascular profiles, I look at proteinurias…"* |

Abbreviations: CHF, congestive heart failure; CKD, chronic kidney disease; DKA, diabetic ketoacidosis; DPP4s, dipeptidyl peptidase-4 inhibitors; GLP1a, glucagon-like peptide 1 analogues; HFrEF, heart failure with reduced ejection fraction; SGLT2i, sodium glucose co-transporter 2 inhibitors; UTIs, urinary tract infections.

[a] Definitions of the parent codes were developed based on Atkins L, Francis J, Islam R, et al. A guide to using the Theoretical Domains Framework of behaviour change to investigate implementation problems. *Impl. Sci.* 2017;12:77.

**Appendix C. Themes and subthemes relating to SGLT2i and GLP1a prescribing, with specific examples and sample quotes.**

| Themes and Subthemes | Definitions | Specific Examples | Sample Quotes |
|---|---|---|---|
| **Barriers** | | | |
| Limited access to medications | Clinicians articulate barriers that reduce patient access to SGLT2i/GLP1a. | | |
| Cost of the medications to patients | Financial considerations limit patients' ability to use SGLT2i/GLP1a, which restricts provider prescribing. | [1] Clinicians care for patients who lack sufficient drug coverage to be able to use these medications, and cannot afford to pay for them out of pocket. This includes patients who are unemployed, under age 65 and not covered by the Ontario Drug Benefit plan, or otherwise facing limited drug coverage.<br><br>[2] Criteria to receive drug coverage exclude some patients who might benefit from the medications. | ***Nurse practitioner:*** *"I'd probably say the biggest issue is cost in our area....we're in a very older-adult, geriatric community, kind of lower income, not a lot of patients with private insurance or benefit plans, so that conversation is basically: let's see what [the Ontario Health Insurance Plan (OHIP)] covers, if anything, and we go from there. And then if there's no OHIP coverage, if they're under 65, we're kind of looking at the next, either least expensive medication on the market that may help us, or is there a compassionate drug program that we can use."*<br><br>***Family physician:*** *"...the other barrier I should mention as well is, now, with the changes in the LU codes for Ozempic specifically, it's just so complicated. Because a lot of patients were on it, like the patients without diabetes but they were on it for weight loss, and now they have to—like if they have diabetes, they have to be on metformin to be on Ozempic and that's when their insurance will cover it. Anyways, that process has been a bit painful because there's a lot of patients who haven't been able to tolerate metformin and they're only on a GLP1, like Ozempic, but their insurance gives them a hard time."* |
| Insufficiency of Compassionate Drug Programs | Clinicians describe challenges using Compassionate Drug Programs that make them an unreliable option for uninsured patients who require treatment with SGLT2i/GLP1a. | [1] Supply of medications is delayed month-to-month.<br><br>[2] Supply of medications terminates after one year. | ***Family physician:*** *"But it's difficult. They take a really long time, and they send a month at a time, and, you know, by the time they finish the first month, we need the next month, right? So I don't find the Jardiance Compassionate Program very effective...we have to fill out lots of forms, and they literally send you one month at a time...And it runs out. Well, yeah, it runs out before they send you the higher dose. Right, 'cause you've got to take the lower dose for one month, then go up. And they're behind the times on sending it...And it takes them 2 or 3 months to send the first month."* |

| | | | | |
|---|---|---|---|---|
| | Periodic drug shortages | Clinicians are limited in prescribing SGLT2i/GLP1a because of their periodic absence from local pharmacies and/or formularies. | [1] Specific medication doses or formulations are absent from hospital and local pharmacies.<br><br>[2] There are periodic GLP1a shortages, lasting up to months. | ***Family + emergency physician:*** *"I think that there's times when certain pharmacies don't carry the dose or the formulation that you're trying for with the GLP1s. I think I've run into that on a couple of occasions, where the electronic medical record that I'm working with at a given clinic will populate a certain dose for the GLP1s and I'll click on that thinking, 'Okay, this is the initial dose. Sounds good. We'll follow up this patient in four to six weeks,' and then the pharmacy will contact me saying 'We don't have that.'"*<br><br>***Resident physician:*** *"There was actually, I think, in Ontario, there was a shortage in general in the springtime slash winter of last year. But even now, in our own hospital, there are at time shortages and even no Ozempic available to the point where patients will have to bring in their own GLP1 if they've already been prescribed it, so that's a main barrier there."* |
| | Clinician shortage | Patient access to SGLT2i/GLP1a is reduced due to limited access to primary care providers who may be able to initiate these medications. | [1] Clinician shortages mean that there are fewer primary care providers available to manage chronic diseases in Northern Ontario and initiate SGLT2i/GLP1a.<br><br>[2] Overworked primary care providers in Northern Ontario may face barriers to incorporating new treatments into routine practice, limiting patient access. | ***Family physician:*** *"...patient education and compliance. But that takes a lot of work in terms of like community resources and education. I mean, like, yeah, we just don't have the – the diabetes care isn't the best 'cause we have like one diabetes nurse. Or there's like, some teams, but on the whole, it's not really—it's quite poorly managed."*<br><br>***Nurse practitioner:*** *"...in northern Ontario, primary care kind of shoulders a lot of it. You know, there's maybe not as readily available consults or they're not followed as regularly by cardiology as maybe as they would be in the inner city. It's not as easy to get these specialities right away, so then it becomes just primary care management. And if, if that's the case, it may just be delayed or kind of not having that speciality up-to-speed ideas...and in Northern Ontario, because of whether its travel for the patient or just available specialities or access, it might be a barrier in like actually getting someone to say: "hey, how about you go put them on an SGLT2 or GLP" and "this is what I would suggest" and "can you do that?" In primary care, just 'cause you're managing so much, it might be a little bit slower to get there, unless there's more in-service or education in those fields...it's more just possibly not thinking of it as much or not seeing possibly the new areas of more specific benefits of that medication and the research. Whereas specialists will probably be more on top of that, like a nephrologist or cardiologist."* |
| Patient Refusal | | Clinicians articulate that their ability to | | |

| | | | | |
|---|---|---|---|---|
| | | prescribe SGLT2i/GLP1a is limited by patients refusing the medications when offered. | | |
| | Medication averse | Clinicians describe experiences where patients refused to take an SGLT2i or a GLP1a out of a general aversion to taking medications or specific aversions to GLP1a. | [1] Patients do not want to increase their pill burden.<br><br>[2] Patients are resistant to injections (GLP1a).<br><br>[3] Patients refuse GLP1a to avoid stigma.<br><br>[4] Patients are resistant to changing medications once stable on other agents. | ***Family physician:*** *"...the patient population I work with is kind of like medication averse, it's really hard to convince them to start so many medications all at once."*<br><br>***Resident physician:*** *"So, the GLP1s, the self-injection with some people that aren't- that are kind of resistant to that."*<br><br>***Resident physician:*** *"I would say 75% of the time to be a rough estimate [of prescribing SGLT2i/GLP1a]. As far as what the barriers would be, like where the other 25% is coming from, I would say patient hesitation sometimes. There's, as you know, these medications are heavily involved in the media. There's a little bit I think of stigma around some GLP1s."*<br><br>***Nurse practitioner:*** *" I think when something's status quo, I don't play with it because patients tend to be resistant to change. Why fix it when it's not broke, sort of thing."* |
| | Fear of side effects | Clinicians describe that patients' fears of and experiences with side effects affect patients' willingness to start or continue on a SGLT2i/GLP1a. | [1] Patients refuse starting SGLT2i/GLP1a after hearing about their side effect profile.<br><br>[2] Patients refuse continuing on SGLT2i/GLP1a after experiencing side effects. | ***Family + emergency physician:*** *"People are sort of not—they don't like the side effect profile. Then, they would prefer to see the alternative options before going down that [GLP1a] route."*<br><br>***Nurse practitioner:*** *"Patients with diabetes are usually obese, so when you start an SGLT2, there's a higher risk of developing fungal infections in the genitalia or the skin folds. And there's a number of patients who have multiple episodes of these, and after 2 or 3, decide that the SGLT2s are not for them, so they stop. That's a major barrier, because I do believe it's a good drug, a good class of medications, but I would say, the recurrent fungal infections are a major barrier for sure."*<br><br>***Nurse practitioner:*** *"And then the GLP1a, I certainly have way more people stop those meds than any others due to the GI side effects."* |
| | Medication non-adherence and appointment no-show | Clinicians are limited in their ability to prescribe SGLT2i/GLP1a for patients who may benefit but struggle with | [1] Some patients struggle to adhere to medication regiments and stop taking SGLT2i/GLP1a without | ***Family physician:*** *"We have a lot of issues with compliance. So, I would say that's the biggest problem. If you ever end up going to North Ontario, and you go to the nursing stations, you will see fridges and fridges and* |

| | | | | |
|---|---|---|---|---|
| | | adhering to medications or committing to appointments. | letting clinicians know. | *boxes and boxes of, I mean, Ozempic, in the fridge that's not used, insulin that's not used. So, the biggest thing is really medication non-compliance. So I would say, we struggle, because you'll see lots of A1Cs that are quite high because a lot of people aren't using those medications...there's social things, right? They'll say "oh, they never call me 'cause of so-and-so." Like, it's actually quite nuanced, I think. There's a lot of mistrust, right? Of like the healthcare system."*<br><br>***Nurse practitioner:*** *"I see complex patients and truthfully, many of them are in that state because they have patient non-compliance or they won't even go to their appointments. That's a big portion of our population, for sure... [The reasons are] kind of all over the place. I would say the biggest one is cost. The next one would be, I think, it depends on if they're on insulin as well, they get a little bit burnt out. And then they get frustrated, and they don't feel like it's helping and then they just want to go off of it. Another one would be, they feel like there could be side effects but they're not sure, and they don't tell the provider, they just go off of it on their own....There's huge amount of no-showing. Like huge. Like, even when we've made therapeutic relationships with them."* |
| Lack of familiarity | | Clinicians identify some uncertainties regarding SGLT2i/GLP1a prescribing that stem from a lack of experience or familiarity with the medication classes. | | |
| | Not enough experience with patients using the medications | Clinicians' uncertainties prescribing or handling SGLT2i/GLP1a arise from relatively limited experience initiating the medications and managing large patient populations who are routinely taking the medications. | [1] Providers struggle to handle adverse drug reactions.<br><br>[2] Providers give greater weight to medication side effects.<br><br>[3] New residents lack exposure to SGLT2i/GLP1a use in certain clinical scenarios. | ***Nurse practitioner: "****With the GLP1s, I'm finding maybe sometimes physicians…family docs are more inclined to… if the individual has any sort of side effect with it, like with the nausea and such, they're a bit more quick to stop them. And it may just be a dose-related complication, and they tolerated the lower dose but not the higher dose, but rather than having the patient reduce to the tolerated dose to still get some benefit, they just stop it altogether."*<br><br>***Family + emergency physician:*** *"I tend to probably overweigh that potential risk [of using a GLP1 just for weight loss], just because I think the alternative of trying to pursue aggressive dietary and lifestyle modifications probably carries far less risk than that....and knowing that sometimes, if I'm doing locum family practice coverage, I'm not sure how closely this patient will be followed up or whether they'll be taken off of* |

| | | | | |
|---|---|---|---|---|
| | | | | *this medication or whether it'll just be renewed for a prolonged period of time and they'll be exposed to risks for longer than what's required or what's needed…"*<br><br>***Nurse practitioner:*** *"Other things, like... I went to a talk recently in Ottawa and it kind of scared me about adding, like, some of the SGLT2s because of risk of diabetic ketoacidosis. So then I got…you know, I guess I have my own nerves."*<br><br>***Resident physician:*** *"Maybe there's a lack of knowledge on my part or just not seeing my preceptors [prescribe SGLT2i for adults without diabetes]—I'm just following suit,"* |
| | Knowledge gaps | Clinicians identify their own knowledge gaps related to SGLT2i/GLP1a prescribing or recognize inappropriate use of SGLT2i/GLP1a among colleagues. | [1] There is a need for greater clarity on when to initiate SGLT2i/GLP1a in patients with chronic kidney disease.<br><br>[2] Some clinicians note their colleagues inappropriately starting SGLT2i for patients with type 1 diabetes.<br><br>[3] There is a need for greater clarity on how to manage patients with severely elevated A1C values (e.g., balancing insulin needs with a desire to initiate SGLT2i).<br><br>[4] Choice paralysis and information overload create uncertainty when selecting a medication for diabetes management in an ever-evolving diabetes drug landscape or even when selecting specific agents within each medication class. | ***Nurse practitioner:*** *"I do [prescribe SGLT2i/GLP1a for patients with chronic kidney disease], although I'm fuzzy on when that should start. So, I know it's a huge indication for it and I'm renewing it often once nephrology or whomever starts it, but I can't say I could give anyone – like a colleague or someone that I'm training a really great explanation on when you would start that."*<br><br>***Resident physician:*** *"...the SGLT2s, the main thing I've seen, which may be more of a provider education issue, is that patients will come in with [diabetic ketoacidosis] and they're on an SGLT2 but they're a type 1 diabetic, and as you may know, that's a big no-no as far as prescribing. So, you know, it's technically a side effect but this patient should've never been on the SGLT2i…and I've actually seen two cases of that in the past two weeks, so that's something that's happening. And again, I think just more of a provider education issue, I think that they'll see that they have heart failure and diabetes, but kind of neglect the diabetes side and put it on for the heart failure management not knowing that it's actually contraindicated for the type of diabetes that they have."*<br><br>***Nurse practitioner:*** *"I'm finding that people are more and more coming into hospital on some of these medications but their glycemic control and their insulin needs are not addressed first and so we're starting them [on SGLT2i] with the hope of the cardiovascular benefit and everything and maybe even just trying to keep them, you know, minimize insulin, but these are individuals who already are at a point in their disease that their insulin—they have insulin deficiency, so that's not being addressed first* |

| | | | | |
|---|---|---|---|---|
| | | | | *and so these are people that unfortunately, I think are having complications probably because that piece is not being addressed and now we just added a layer to things that are kind of making the symptoms worse."*<br><br>***Nurse practitioner: "****I think the biggest thing is the education, knowledge aspect of things. You know, I remember when I was going through school, treating type 2 diabetes, it was either metformin or gliclazide, and if those didn't work, insulin. And it was easy. And now there are just so many more diabetic drugs out there and things are, you know, it's a changing landscape all the time. So I think it's just the knowledge and getting access to the education piece that goes along with these is probably the biggest thing that will help us, or the lack of knowledge, I guess, or confidence or experience is the biggest barrier."*<br><br>***Resident physician:*** *"...on UpToDate, I was trying to read between them, like which one's better than the other, and it's hard to tell....there's too much knowledge, and when you're reading through all of it, they're telling you that they all tested the same in a way, you know, so then you're trying to figure out, like ok, is one better...can you use one over the other for like someone who has heart failure versus kidney failure versus like both. So there's no deciphering answer out there for you. You kind of just have to pick the best one and go with it, so I just went with what's familiar to me."* |
| | Lack of confidence | Clinicians describe lower confidence in their skill handling SGLT2i/GLP1a. | [1] Providers identify challenges specific to handling SGLT2i/GLP1a in patient care (e.g., dosing, titration schedules, holding the medications around procedures, etc) | ***Nurse practitioner:*** *"I talk to my colleagues about a lot, is with the SGLT2 inhibitors, is amputation risk. Because we're in hospital settings, we have quite a few patients that are having acute amputations and stuff like that, so I do discuss that pretty frequently about like, when I should be holding this, should I still start this, should I not start this?"* |
| Clinical identity | | Clinicians' SGLT2i/GLP1a prescribing behaviours are influenced by their personal beliefs and perceived clinical role. | | |
| | Risk tolerance | Clinicians describe being hesitant to prescribe SGLT2i/GLP1a for patients for whom they believe the risks to outweigh the benefits. | [1] Some clinicians prefer to delay prescribing SGLT2i/GLP1a in:<br>(i) Unstable hospitalized patients until they have been stabilized or upon discharge | ***Family physician:*** *"...if they are coming in with like, diabetes and heart failure, you know, like, let's try to get them out of the acute decompensation phase of the heart failure, and once they are, then we can kind of start, like, with starting medications. It would kind of be like a step-wise approach because of the patient population that I have to deal with."* |

| | | | | |
|---|---|---|---|---|
| | | | (ii) Patients with severely elevated A1c until they are first stabilized on insulin<br><br>[2] Some clinicians prefer not to prescribe SGLT2i/GLP1a at all for:<br>(i) Patients who are not obese<br>(ii) People taking several diuretics<br>(iii) People who are really ill (i.e., have gastroparesis, pancreatitis, amputations)<br>(iv) Patients for whom lifestyle interventions may work<br>(v) Patients who many not seek medical attention if they are unwell<br>(vi) Patients with diabetes who are stable on other anti-hyperglycemic agents<br><br>[3] Clinicians find the oral semaglutide formulation to have bad poor side effects and poor efficacy.<br><br>[4] Clinicians fear renal insult with SGLT2i. | ***Nurse practitioner**: "For SGLT2 inhibitors, if an individual is elderly with mobility and/or cognitive issues, I am finding that because I am and… and perhaps their glycemic control is not really optimal just yet so they're already experiencing some glucosuria and frequency because of that, I would probably avoid an SGLT2 inhibitor in that scenario. People who've come in with frequent urinary tract infections and cog—and again, probably elderly with cognitive issues—I would probably steer clear in that scenario as well."*<br><br>***Family physician:** "Empagliflozin I- I generally prefer, I get nervous putting women on it because they all get bladder infections and yeast infections, so… so I-I, but again, in choosing between them, I do involve the patient in the decision-making process."*<br><br>***Nurse practitioner:** "If they're stable, I tend not to change. There's no hypoglycemic episodes, A1C stable, I leave them on it."*<br><br>***Nurse practitioner:** "I have very little issues with that. But that's where we need the research though, because a lot of physicians have issues with that. 'Cause they think it leads to hypovolemia and it leads to worsening renal function, and they can't handle the dip, because there's always a little bit of a dip when you start the SGLT2s as well in the eGFR and creatinine. So, I'm ok with that and I have read a lot about it and I've seen talks about it, and I feel really confident with it, but we do have to spend a lot of time educating others about that because they get very upset."* |
| | Perceived scope of practice | Clinicians believe that prescribing SGLT2i and/or GLP1a falls outside of their clinical responsibility. | [1] Primary care practitioners believe it within the domain of nephrologists to prescribe SGLT2i for patients with chronic kidney disease.<br><br>[2] Emergency care physicians do not wish to prescribe SGLT2i/GLP1a ahead of primary care providers and have limited capacity for follow-up. | ***Family physician:** "I don't manage patients with chronic kidney disease."*<br><br>***Family + emergency physician:** "I would say it's a bit different in my situation when I'm working in the emergency department for these patients that tend to have a little bit of a different lens on things that way, like where for some patients more than others, their primary care physician I usually tend to try to not step on toes and I wouldn't tend to introduce things in the emergency department, unless the patient doesn't have a primary care provider or adequate follow-up and it was a primary reason for the presentation, for example."* |

| | | | | |
|---|---|---|---|---|
| | Hesitancy accepting GLP1a for weight loss | Some clinicians' personal beliefs hold them back from considering GLP1a earlier in the care of those who may benefit from weight loss or expressly request to be started on semaglutide. | [1] Some clinicians advocate for longer trials of lifestyle modifications before considering GLP1a for weight loss, expressing less comfort with using GLP1a for this indication.<br><br>[2] Some clinicians describe experiences with colleagues who do not consider obesity a chronic disease that should be treated pharmacologically. | ***Resident physician***: *"I think like for obesity, the first line should always be, kind of like, lifestyle and talking to them about diet and seeing if there is a dietician available...I think like especially now with the commercials about GLP1s, more and more people are asking about them just specifically for obesity management even regardless of their status of like diabetes. So, I feel like the conversation is just becoming more frequent for patients—especially in primary care, when I did family medicine in med school—patients are coming in and asking for them. But, right now, like, if I saw a patient in front of me, where let's say, if they are obese but they don't have diabetes, then I probably wouldn't necessarily think to just like jump on a GLP1. Like, I feel like a lot of my preceptors and my comfort right now is not necessarily to use that as first line."*<br><br>***Resident physician:*** *"I'll go back to the prescription of Ozempic even off-label, I mean right now, I guess they're working on using these medications just for weight loss itself, but I feel like there's a lot of pre-conceived notions or pre-conceived thoughts on just obesity in general. With certain doctors, I just remember listening to a talk from one of the residents, so it was like residents teaching residents, and she was like this super active concern, she was like 'don't even get me started on patients wanting Ozempic that are obese' and I was like, I don't think this is the approach that you should take. This is just a personal choice, I guess, because even if they do bariatric surgery or a sleeve surgery, it's almost the same thing. There is some sort of psychological aspect or psycho-social aspect with obesity as well. But if you are seeing numbers, like they are now pre-diabetes, I would think as a physician, practicing more preventative medicine would be better than just saying, you know, that they're going to put back on the weight. I feel like there's some disconnect in the doctors viewing patients in that way."* |
| | Clinical practice guideline-directed | Clinicians' adherence to clinical practice guidelines can conflict with their expectations of the benefits of using SGLT2i and GLP1a. | [1] Clinicians felt obligated to follow clinical guidelines and prescribe metformin as the first-line agent in diabetes management, even when they believe that patients may benefit from an earlier SGLT2i/GLP1a prescription.. | ***Family physician:*** *"I know [the guidelines recommend metformin as first line], but the question is why do they? It's probably cost or just history, but they still do. But that might change, I guess, over time."*<br><br>***Nurse practitioner:*** *"...oftentimes, I'll be more comfortable with the SGLT2s in those patients. So, looking at, you know, if they have a diagnosis of diabetes, and going through the guidelines, often starting with metformin, but sometimes that is a barrier, because if they have heart failure or are at risk for that or need some renal protection, would prefer to* |

| | | | |
|---|---|---|---|
| | | | *almost straight away or lean on an SGTL2 because of the benefits that they have in heart failure and renal protection."*<br><br>***Nurse practitioner:*** *"Yeah, I've seen that starting to become more of a trend, even in consult notes and stuff, to just try and go straight to [SGLT2i/GLP1a]. It still seems like metformin hangs around a lot, like, you know, it's an obligation to start on it before going to an SGLT2, however, there's getting more comfort on trying to go straight to those."* |
| Prescribing inertia | Clinicians refer to their routine prescribing behaviours or algorithms that they must change to incorporate SGLT2i/GLP1a. | | |
| Routine patterns of prescribing | Clinicians allude to an adjustment period required to translate new knowledge of SGLT2i/GLP1a into new prescribing habits. | [1] Clinicians describe forgetting to counsel or prescribe SGLT2i/GLP1a as they are building new prescribing routines. | ***Nurse practitioner:*** *"Yeah, I think it's just an omission on my part, and if I think about it, because I did read the last study on it, which was actually very compelling. I guess it just hasn't translated yet into practice for me."*<br><br>***Resident physician:*** *"...maybe it's like closer to 50-50 right now, where we're remembering to bring it up with patients. Trying to…*" |
| **Facilitators** | | | |
| Role as a clinician that follows the data | Clinicians describe the data they consult when making prescribing decisions, which is either tied to their role as a clinician and more personally tied to their identity as a clinician. | | |
| Respecting clinical guidelines | Clinicians describe making prescribing decisions based on guidelines. | [1] Clinicians prescribe SGLT2i/GLP1a based on recommendations included on Canadian clinical practice guidelines, including Diabetes Canada, the Canadian Cardiovascular Society (CCS), and Kidney Disease: Improving Global Outcomes guidelines (KDIGO). | ***Resident physician:*** *"...especially SGLT2s are like kind of one of the four pillars of HFrEF management now. So, in, even like outpatient internal medicine when I did some rotations last year, and inpatient now, I've definitely seen, like, we're thinking about SGLT2s more and even just within a few years of me doing, like, rotations in inpatient medicine, I find, like, during rounds, if we have a patient with diabetes especially and HFrEF, then everyone's like trying to see, "oh, are they on an SGLT2?" and we're making sure that we're covering that pillar of heart failure management. So I think it's becoming more talked about and more common that we're prescribing them for that."* |

| | | | | |
|---|---|---|---|---|
| | | | | ***Family + emergency physician:*** *"[I prescribe] usually going by Diabetes Canada guidelines and recommendations."* |
| | Guided by evidence | Clinicians describe making prescribing decisions based on emerging evidence. | [1] Clinicians prescribe SGLT2i/GLP1a based on compelling evidence of their benefits in recent clinical trials. | ***Family physician:*** *"I try my best to make sure that what I'm doing is evidence based."*<br><br>***Family physician:*** *"...in general, we have switched first line—until about a year ago, we thought of first line as being metformin—now we look more to empagliflozin or Ozempic, semaglutide I should say, depending on the patient's needs and other factors with the patient. And drug plans! [I refer to] Trials, more so. 'Cause the empagliflozin trials seem to have the most cardiac protection than any of them. And, whereas metformin, you go way back to UKPDS and there's been a number of other studies that show it has a slight improvement in mortality, but it doesn't seem to have the same strength as the other two."* |
| Belief that SGLT2i/GLP1a will improve patient outcomes | | Clinicians articulate beliefs that SGLT2i/GLP1a use will improve patient health. | | |
| | Desire to offer their patients medications with strong benefit profiles | Clinicians express an explicit desire or motivation to prescribe SGLT2i/GLP1a because the medications offer benefits beyond glycemic control. | [1] Clinicians are motivated to prescribe SGLT2i/GLP1a to offer their patients cardiovascular and renal protection and/or help with weight loss.<br><br>[2] Clinicians are motivated to prescribe SGLT2i/GLP1a to reduce patients' risks of mortality.<br><br>[3] Clinicians are motivated to prescribe GLP1a because they have been shown to reduce insulin. resistance.<br><br>[4] Clinicians are motivated to prescribe SGLT2i/GLP1a because they believe these agents will | ***Nurse practitioner:*** *"...metformin is generally the first pick. However, considering most patients have an issue with weight, and also that there's a class of medications—well multiple now—but there is a class of medication that is now known to decrease further the risk of cardiovascular complications, I do try to favour Synjardy, which is a combination of metformin and an SGLT2 inhibitor. So that's usually my first step for most patients, and also considering the fact that it can lead to a small weight loss, which is always appreciated."*<br><br>***Resident physician:*** *"...just the fact that they can decrease mortality. And it's been proven. And it's, you know, just-just that alone is enough for me to want to put a patient in heart failure on it. Let alone having diabetes and wanting to protect their kidneys as well, I know it's protective against diabetic---like—renal nephropathy. There's that benefit as well....So that's my go-to for standard of practice right now."*<br><br>***Nurse practitioner:*** *"It's kind of new to me to start someone initially, as a primary- as the first medication of choice to be one of the GLP1 receptor* |

| | | | | |
|---|---|---|---|---|
| | | | generally improve patients' quality of life or meaningfully impact health outcomes. | *agonists, Ozempic and such. But we have the discretion as a group of healthcare providers, and we think we believe it's pretty acceptable to do that. My concern was that I really like...tackling the insulin resistance thing. And now I'm understanding that something such as Ozempic...helps with that insulin resistance piece, so I'm getting comfortable prescribing it as a first line, especially in patients who are struggling with obesity."*<br><br>***Nurse practitioner:*** *"...if there's a medication that can help with the diabetes but also patients' quality of life, help them with that, then it's definitely something we're interested in and would start the patient on if they're interested in starting it."* |
| | Optimism that medication use will improve health outcomes | Clinicians have positive perceptions of the ability of SGLT2i/GLP1a to improve the health outcomes of patients. | [1] Clinicians express optimism about long-term SGLT2i/GLP1a use based on knowledge of clinical trial data.<br><br>[2] Clinicians express optimism about short-term SGLT2i/GLP1a efficacy based on experience with patients having good outcomes. | ***Resident physician:*** "*I always tell people, even my family, even if I don't have high blood pressure or diabetes, I always want to start myself on an ACE inhibitor in my 60s just because it's cardioprotective and renal protective. But I also think the same thing about the SGLT2 inhibitors, as well!"*<br><br>***Family + emergency physician:*** *"I think in comparison, especially to some of the other second-line medications for type two diabetes, I think they hold excellent value for reducing long-term morbidity and mortality, especially as it relates to cardiovascular disease. I think more so the SGLT2s related to CHF, but the GLP1s, I think also have some research related to cardiovascular disease outcomes and even renovascular disease outcomes. Like, reno- microvascular disease. I think, as far as reducing the declines in GFR and reducing the risk of at least morbidity from cardiovascular disease, and I think a couple studies have even pointed towards reducing mortality from cardiovascular disease, for both the SGLT2s and GLP1s if I'm not mistaken. So, I think those findings alone are something to be excited about but something to sort of push for in this population when it's indicated."*<br><br>***Nurse practitioner:*** *"...when the patient is using them correctly, I do see great improvement. It reinforces their behaviour, and it reinforces mine."*<br><br>***Nurse practitioner:*** *"I'm noticing patients with—who are able to tolerate them and were able to continue. They are living longer with heart failure, I think. I'm seeing those with—and retaining functional and meaningful capability, as well. Patients are having delayed progression of renal* |

| | | | | |
|---|---|---|---|---|
| | | | | *disease and I am noticing that as well, so we're having—I don't have an exact statistic or a number, but I have recently been told that, within the last year, I believe, Sudbury has seen the first decline…or maybe, not decline, but not an increase in the number of diabetic patients going on dialysis."* |
| Clinicians' perceptions of patient openness | | Clinicians describe patient-level factors that they perceive to facilitate SGLT2i/GLP1a prescribing. | | |
| | Patients express desire for weight loss | Clinicians note that GLP1a prescribing is facilitated by patients who come into appointments specifically asking to be started on semaglutide. | [1] Some patients looking to lose weight tend to choose a GLP1a agent when provided with different treatment options or prompt GLP1a prescribing earlier in their care via explicit request. | ***Nurse practitioner***: *"Patients are well aware that the extent of coverage is greater when diabetes is a known diagnosis. Right? So I have to say, most of my overweight patients who for some reason do become diabetic, they already know that they are eligible for Ozempic. So, in my office, they'll ask me: "XX, I want Ozempic." The other scenario is I have an obese diabetic in my exam room and I know that they have a very good entrance coverage, and [in] my plan section of my encounter, I will discuss the different options and I will explain the benefits of Ozempic and of course the risks. And usually, by that point, because they generally want the weight loss, they will also select to go with the Ozempic."*<br><br>***Nurse practitioner:*** *"...for patients who are struggling with weight and weight loss, I mean, the GLP1s, mainly Ozempic because it's been so popular in the media, is always a part of that discussion to see if weight reduction can be done through those. I know SGTL2s can do that a little bit, but more just probably because of the awareness of it, we end up discussing the GLP1s a lot more."* |
| | Patients understand second-line treatment may be needed | Clinicians perceive that some patients may be more willing to start on an SGLT2i/GLP1a if they are made aware that another medication may be needed later in their care, which facilitates prescribing when these agents are later indicated. | [1] Patients who are introduced earlier in their care to the potential need for second-line therapy to manage their diabetes may be more willing to take on an SGLT2i/GLP1a. | ***Family physician:*** *"...when we're diagnosing this, there's a lot of information, but, you know, once you get into, kind of, the 3 months, 6 months, even patients will know that their A1C's been creeping up. We'll be like: "You know, your A1C used to be really good. It used to be 6.5, then it's at 6.8, now it's 6.9. Now, it's 7. The trend for the past 2 years has been that it's going up, we're probably going to have to add another medication. I think if we start talking about that earlier, it makes it easier when it's like, 'Listen, your A1C is now 7.5 for the second 3-month check-in in a row. It's time to try something else.' It's not the first time they're hearing that. They've had, you know, a few months to digest that. Then they're often more prepared and willing."* |

| | | | | |
|---|---|---|---|---|
| Comfort Prescribing | | Clinicians identify several factors that increase comfort and confidence prescribing SGLT2i/GLP1a. | | |
| | Experience builds skill and confidence | Clinicians describe that experience counselling, prescribing, and managing the use of SGLT2i/GLP1a for patients with diverse medical profiles enhances their confidence with these medications. | [1] Clinicians feel more confident prescribing because they see good effects in the patients they start on SGLT2i/GLP1a.<br><br>[2] Experience prescribing helps build skill and confidence managing side effects and handling the medications (e.g., dosing, titrations). | ***Resident physician:*** *"I would say SGLT2, reasonably confident. I've done it a lot of times. Seen good effect."*<br><br>***Nurse practitioner:*** *"...having had 30% of patients who do not tolerate GLP1 agonists, having to manage that. So, made me more confident. So initially when they had these severe symptoms, I was a bit unsure of what to do, but I've had a number of them now, so I feel more confident and competent, so experience for sure.* |
| | Prevalence of SGLT2i/GLP1a builds comfort prescribing | Clinicians nod to the increasing prevalence of SGLT2i/GLP1a use as a factor that helps normalize prescribing SGLT2i/GLP1a. | [1] Noticing colleagues prescribe or discuss SGLT2i/GLP1a more often helps clinicians feel more comfortable prescribing the medications themselves.<br><br>[2] Widespread marketing of semaglutide fosters the perception that these medications are common and broadly accepted. | ***Resident physician:*** *"...like I think in the last few years especially, it's something that's regularly coming up in discussion and even like on discharge planning, I hear my colleagues—like, my other residents and staff physicians—talking about making sure that someone's on a GLP—or an SGLT2 when they're leaving hospital with heart failure. But I think it's yeah, it's becoming more common."*<br><br>***Family + emergency physician:*** *"...seeing more and more folks on them in the emerg department setting, and then, sort of, reading more about it and then just prescribing it more...they just seem like they've become more commonplace."*<br><br>***Resident physician:*** *"I feel like they are becoming more mainstream. A lot of people know more about them, like I was watching the Olympics and there was an Ozempic commercial like every 5 minutes. So not only are we learning about this as healthcare providers, but the public is too. So, I think that that sets me up for success, basically, with these medications and like, knowing them so well, that when I'm done my residency, I'll be very familiar, very comfortable with them."* |
| | SGLT2i are easy to prescribe | Clinicians describe that initiating patients on an SGLT2i is not mentally or technically onerous, facilitating prescribing. | [1] SGLT2i were approved under patients' Non-Insured Health Benefits (NIHB) before GLP1a, which makes them easier to initiate for patients. | ***Family physician:*** *"...it kind of came up and was approved for coverage with my patients' non-insured health benefits a long time before the GLP1s were, so I was really choosing that over a lot of other choices because it was covered and it wasn't a needle."* |

| | | | | |
|---|---|---|---|---|
| | | | [2] SGLT2i are oral pills with few dosing options, easy titration schedules, and no renal dose adjustments. | ***Nurse practitioner:*** *"So it's very straightforward in a medication like Jardiance, to say ok well, it has some cardioprotective, renal protective features, if individuals are at risk for those or have those especially, then kind of leaning on that, it's pretty straightforward to start the 10 mg and increase to 25 if their target's not met or if they need more and tolerating it well."* |
| | SGLT2i/GLP1a are backed by good data | Clinicians describe feeling comfortable prescribing SGLT2i/GLP1a because they know the medications have been proven to show health benefits in recent clinical trials. | [1] Clinicians directly link comfort or confidence prescribing with their knowledge of clinical trial data. | ***Resident physician:*** *"But I think for like the HFrEF trials with SGLT2s, like, those are quite strong trials so I'd be comfortable prescribing."*<br><br>***Family physician:*** *"Pretty confident [prescribing SGLT2i/GLP1a]. Based on a review of the literature and many recent comments about the guidelines, which—the guidelines, I never thought have been really evidence-based, frankly."* |
| | Prescribing is important to scope of practice | Clinicians feel motivated to develop comfort prescribing SGLT2i/GLP1a because they view it as part of their clinical responsibility. | [1] Clinicians link prescribing to their professional role and patient population. | ***Nurse practitioner:*** *"...definitely, there is a comfort in primary care in starting them and continuing them."*<br><br>***Resident physician:*** *"...the standard teaching is to start with metformin. I think in my experience, I'm always in an inpatient setting and what I would say is I'm much more inclined to be treating the comorbidities, namely the cardiac and CKD."* |
| System and Colleague Supports | | Clinicians describe how system-level facilitators and support from colleagues have enabled SGLT2i/GLP1a prescribing. | | |
| | Expanding eligibility criteria for drug coverage | Clinicians describe that expanding eligibility criteria for drug coverage has facilitated their prescribing. | [1] The removal of financial restrictions enables prescribing. | ***Family physician:*** *"I think the biggest thing is that they have become so much more accessible in the last few years. I remember before, you know, they first came out 5 years ago, that you could not get people on these unless you met all this criteria; you've tried this, this, and that. Like, glyburide and gliclazide, which nobody should be on, right? It made care very onerous, and I think it's overall positive that the, you know, eligibility has been expanded to keep up with best practice and best evidence."*<br><br>***Family physician:*** *"I believe before, [the Non-Insured Health Benefits criteria] required you to fail a glicazide, so you had to be—or sorry, like a – yeah glicazide, or sulfonylurea, you had to fail that. But I think, and* |

| | | | | |
|---|---|---|---|---|
| | | | | *again, I haven't had any issues with it, that now they cover it without needing to fail the sulf- like, that class- that drug class. Which is good. That was a big restriction before."* |
| | Colleagues motivate ongoing learning | Clinicians describe that their prescribing practices regarding SGLT2i/GLP1a have been influenced by the knowledge they acquire about these medications from colleagues. | [1] Preceptors help shape residents' SGLT2i/GLP1a prescribing practices.<br><br>[2] Specialists help shape clinicians' SGLT2i/GLP1a prescribing practices, including internal medicine colleagues, cardiologists, nephrologists.<br><br>[3] Other colleagues help clinicians learn about SGLT2i/GLP1a prescribing. | ***Resident physician:*** *"I even find my attending physicians are trying to teach more around as well to raise awareness and get people well-versed in managing and prescribing these medications...In internal medicine residency, you're getting teaching a lot and you're really advised to stay up-to-date with the literature and the clinical trials."*<br><br>***Resident physician:*** *"...as I've gone through my training as a medical student and now as a resident, there's a lot of experience you've seen from preceptors that they've given advice for, or have had patient success stories or patient not-success stories, on certain medications, and so…from a learner's perspective, definitely hearing perspectives from my preceptors or other colleagues of mine, sharing what their experiences were is definitely beneficial for us. Like, we learned more about metformin and insulin going through medical school, not—I honestly don't think that in my first 2 years of medical school that we really talked about the GLP1s or SGLT2s, until I got into clerkship, which is third and fourth year of medical school. And, there I was really—it really opened my eyes to the benefits of both those treatment options. And not just for diabetes, but literally other things too."*<br><br>***Family + emergency physician:*** *"I tend to lean with those with kidney disease or coronary artery disease towards SGLT2s, but that is—I think that's more of a, like…what would I call that…that's less maybe guideline, and more expert opinion that I—from my colleagues....from the lens of me as an emerg doc or a family doc in the community that I had, I had sort of sought the opinion of my internal medicine colleagues, and they have sort of helped to bridge my knowledge gaps to, sort of, help patient care."*<br><br>***Nurse practitioner:*** *"I guess, another way that I'd gather that information would be from consult reports back from specialists, so whether they be cardiology or nephrology—would be big ones. With their recommendations or suggestions."*<br><br>***Nurse practitioners:*** *"I think it's actually a nurse practitioner from down south who gave the boot camp on diabetes management. It was with her and her nurse, I used to email them back and forth for guidance and I* |

| | | | | |
|---|---|---|---|---|
| | | | | *found them so helpful. And thankfully, I stopped needing to email them. That's pretty much what put me on those two medications"* |

Abbreviations: ACE inhibitor; angiotensin converting enzyme inhibitor; CHF, congestive heart failure; CKD, chronic kidney disease; eGFR, estimated glomerular filtration rate; GLP1a, glucagon-like peptide 1 analogues; HFrEF; heart failure with reduced ejection fraction; LU does, Limited Use codes; OHIP, Ontario Health Insurance Plan; SGLT2i, sodium glucose co-transporter 2 inhibitors; UKPDS, United Kingdom Prospective Diabetes Study.
